# Effects of APOE e4 and Neuropathological Diagnoses on Neuropsychiatric Symptoms: Mediation Analyses and Likely Causation in an Integrated NACC Database

**DOI:** 10.1101/2024.01.30.24301966

**Authors:** Terry E. Goldberg, D.P. Devanand, Zhiqian Fang, Hyun Kim, Elizabeth Rueppel, Aren Tucker, Scott Carlson, Seonjoo Lee

## Abstract

**Background:** Our goal in this study was to identify paths from APOE e4 to neurobehaviors itemized on a neuropsychiatric inventory that involved neuropathologies associated with e4 (amyloid, tau, cerebral amyloid angiopathy, and Lewy bodies) or cognition mediators (memory or global cognitive status), as well as direct paths from e4 to cognition or neurobehaviors.

**Methods:** A total of 1199 cases with available neurobehavioral, cognition and neuropathological data were included. We then conducted a series of causal mediation analyses in R in which e4 always served as the independent variable and Neuropsychiatric Inventory (NPI) neurobehavioral items, when included in the mediation, the outcome. Neuropathologies or cognition served as mediators.

**Results:** Multiple significant indirect paths from e4 through neuropathologies to neurobehaviors were identified. More refined analyses indicated that neuritic plaques and Braak stage, but not extent of diffuse amyloid plaques, drove the findings. A significant direct effect of e4 to memory was also identified. Additionally, Lewy body disease, when treated as an exposure, had a direct effect on hallucinations in keeping with known features of the disease.

**Conclusions:** We found strong evidence for partial mediation of NPI symptoms by cognition, suggesting that cognitive limitations that may have influenced understanding (or misunderstanding) the environment with impacts on maladaptive behavior. In addition, neuritic amyloid plaque levels and Braak stage, but not diffuse amyloid plaque extent, were key in NPI mediated associations suggesting the possibility that synaptic failure play an important role in multiple neurobehavioral symptoms in dementia, including psychosis. Last, we found strong evidence that e4 may have direct effects on cognition when we used verbal episodic memory as an outcome, suggesting that medial temporal regions that support memory may be sensitive to non-amyloidogenic and non-tau related pathophysiological processes.

## Introduction

The Apolipoprotein E (APOE) e4 allele is a known, strong genetic risk variant for Alzheimer’s disease (AD) and AD related dementia (ADRD) histopathologies, including amyloid plaques, tau paired helical filaments (PHF), Lewy body inclusions (LBD) when involvement includes limbic allocortex and neocortex, and the vascular pathology of cerebral amyloid angiopathy (CAA), which involves amyloid deposits on the walls of cerebral arteries and capillaries. Recent work using a variety of methods and models (human imaging, post-mortem brain tissue, mice, and iPSCs) has highlighted that e4 may also be associated with neurobiological compromises that are independent of these pathologies, including lipid production and transport, bioenergetics, involving attenuated mitochondrial function and glucose metabolism, microglial and astrocytic dysregulation, and BBB loss of integrity^1–6^.

It has been known since the late 1990s that e4 increases the risk of neuropathologically verified AD^7^. A large and recent of over 5000 brains demonstrated an OR=6.1 for one copy of e4 and OR=31.2 for two copies when contrasted with e3 homozygotes^8^. E4 also increases the risk of AD when compared to the neuroprotective APOE variant e2^9^. It is also established that E4 is associated with autopsy verified CAA^10–12^. Additionally, e4 has now been associated with Lewy body pathology independent of AD pathologies, especially when such pathology inclusions are found beyond the midbrain in limbic areas such as the amygdala and neocortex.^13, 14^. Goldberg et al showed that the association of e4 and midbrain only LB was negligible, but increased significantly when such pathology spread beyond the midbrain^9^.

The clinical manifestations of neuropathological subtypes of dementia include decline in cognition, decline in function, and neurobehavioral changes. In these neuropathologies, the gross relationship between e4 and cognitive and functional decline characteristic of dementia has been examined, but the impact of e4 on neurobehavioral or neuropsychiatric abnormalities in neuropathologically diagnosed dementia subtypes has not been well characterized.

Our intent in this study was to identify both paths from e4 to NPI neurobehaviors that involved ADRD neuropathologies or cognition mediators or direct paths that did not require mediators. We implemented this approach in a logical and neurobiologically plausible sequence of causal mediation analyses as shown schematically in Figure 1. We hypothesized that e4 may have mediated effects on cognition or neurobehavior via e4 associated ADRD pathologies that increase aggregation of beta-amyloid, tau, and/or alpha-synuclein, i.e., through the indirect path in mediation models. We further hypothesized that non ADRD related neurophysiologic compromises would result in direct path effects of e4 on cognition or neuropsychiatric symptoms, as they would *not* be captured by the multiple neuropathologies (AD, LBD, CAA) under examination. We conducted the study in the NACC Neuropathology and Unified Data Set (N=1199) and integrated genetic, neuropathological, and cognitive data^15, 16^.

**Figure 1.**
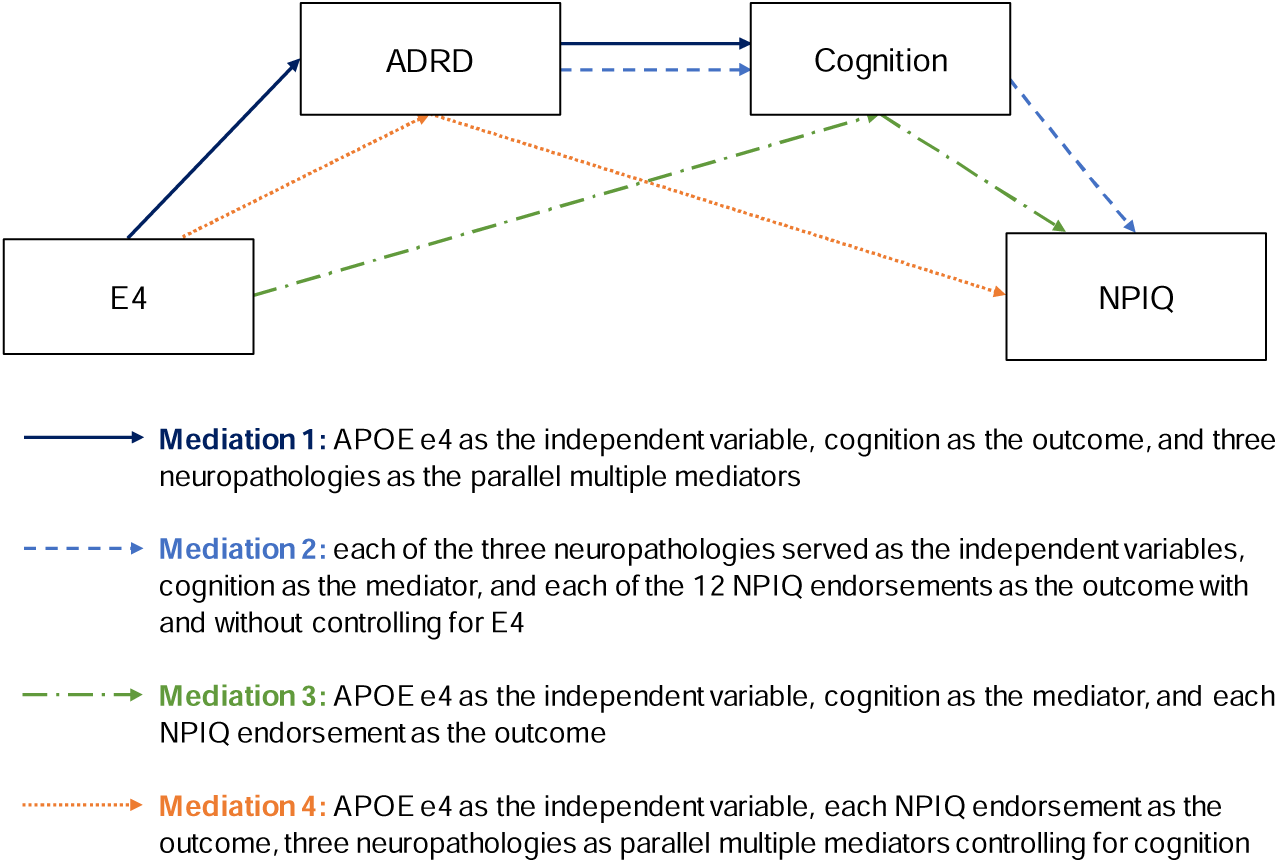
Mediation analyses testing the causal pathways depicted in the schematic figure. In these pathways, e4 (when present) always served as the independent predictor (exposure) and NPI items (when present) always served as the outcome. Histopathology served either as a mediator, or when e4 was controlled, as an exposure. Cognition served either as an outcome (if histopathology was present) or a mediator when NPI items were the outcome. This set of analyses respects the neurobiologic plausibility and logic that e4 is causative for ADRD pathologies (AD, LBD, CAA), which then may cause or otherwise mediate cognitive changes and/or NPI changes.

## Methods and Materials

### Study Sample

After exclusion of the neuropathological diagnoses of brain injury, CNS neoplasm, Down syndrome, Huntington’s disease, and prion disease, we included all brains for which the NPI-Q was administered to a caregiver at least once during life, ApoE4 genotype data were collected, ADRD (AD, LBD, CAA) neuropathological diagnoses were complete, and Logical Memory or Craft Story and MMSE or MOCA and were administered at the last NPI-Q visit. Supplementary Figure 1 shows a flow chart of the sample derivation.

### Genotype

The e4 group combined e3/e4, e4/e4, and e2/e4 genotypes. The latter genotype was shown by us to have risk ORs similar to e3/e4 in this database and has been replicated by others^8, 9, 17^. All other genotypes were in the non-e4 group. <u>Pathologies</u>

We elected to examine APOE genotype associations with the following classes of neuropathologies. Using Montine ABC criteria^18^ a cut off indicating intermediate or high probability of AD pathology in the AD neuropathological change score (ADNC) was used to indicate AD severity in binary analyses. ADNC score was defined by a combination of these three pathologies (diffuse plaques, Braak stage, neuritic plaques) and its score range was 0-3. A score of 2 or 3 (intermediate or high) was considered consistent with AD dementia. The ADNC score is termed AD pathology for simplicity throughout.

1. AD histopathologies were as defined in the ABC criteria for severity. A. Diffuse amyloid plaque (Thal stage) is a measure of spread of plaque and higher scores indicate greater spread of pathology (score range 0-4). B. Braak stage is a measure of progression of NFTs, and higher-level stages indicate spread of pathology to neocortex (0-6). C. Neuritic plaques are a hallmark feature of AD and may have more specificity to AD than diffuse plaques, with higher levels indicating greater density of pathology (score range 0-3).
2. Lewy body disease due to alpha synuclein aggregations, as it has recently been established that there is increased co-morbidity between AD and Lewy body disease and that e4 may promote Lewy body pathology independent of AD pathologies. Ratings were based on midbrain only, limbic, and neocortical involvement (score range 0-3). E4 is associated with limbic and neocortical alpha synuclein aggregates in the NACC sample. It was rated as present/absent.
3. CAA, characterized by the accumulation of amyloid beta in the walls of small to medium sized vessels, is associated with APOE e4 in the NACC sample. Ratings were based on severity of the deposition (0-3). It was rated as present or absent.

We accessed the NACC Neuropathology Data Set v. 10 (December 2016) for this study. This version assembles neuropathological data collected in a uniform manner from 39 Alzheimer’s Disease Center (ADC) sites in the United States^15^. Semi-quantitative ratings were made by immunohistochemistry, histochemistry, microscopic visualization, or visual inspection and appropriate regional examinations. We merged the neuropathology database with the NACC Unified Data Set of clinical longitudinal data^16^. Our sample included 1199 cases with APOE, memory (delayed recall), neuropathological and NPI data. We included all cases with such data irrespective of clinical diagnosis. As such our study was transdiagnostic and thus not susceptible to clinical misdiagnosis.

Severity ratings for each pathology are described in detail in the NP Codebook. In all cases 0 indicates absence of pathology and higher numbers indicate more severe pathology. For ADNC 0 and 1 were considered absent and 2 and 3 were considered present. For other binary analyses pathologies were rated as absent/present. For ordinal analyses in the ABC criteria, we used the entire score range for the pathologies.

### Cognition

Logical Memory II (delayed paragraph recall). This is a test of verbal episodic memory involving consolidation of recently acquired material after a delay (range 0-25). Failures in consolidation, usually evident in delayed recall, are considered a cardinal feature of AD. When this measure was unavailable but another memory for stories measure, the Craft Story, was available (N=112), we converted the latter into a Logical Memory II score using the formula provided in the NACC crosswalk study^19^. Scores coincided with the last NPI visit.

MMSE, used in sensitivity analyses, is a screening test of global cognitive status (range 0-30 points). For MOCA results (N=139) we converted scores to MMSE scores using a NACC crosswalk table^19^.

### Neuropsychiatric Inventory (NPI-Q)

We examined the 12 items of the NPI-Q, hereafter called the NPI^20^. The NPI measures severity of neurobehaviors on a 0-3 scale and is based on an informant interview. We binarized each item on the basis of present (a rating >0) vs. absent and used the NPI final assessment prior to study withdrawal or death.

### Covariates

For all analysis, we included age at death, sex, education and time in years between last available NPI assessment and death as covariates.

### Statistical Analyses

Statistical analysis was performed using R version 4.1.1.1 For descriptive statistics, means and SDs and frequency and percentage were reported for continuous and categorical variables, respectively^21^. For statistical testing, Pearson’s chi-squared test and Kruskal-Wallis rank sum test were performed as appropriate.

We first tested the association between e4 and the three ADRD pathologies using logistic regression, controlling for the covariates. The odds ratios (ORs) of e4, 95% confidence intervals and p-values were reported.

We hypothesized that the association between APOE e4 and NPI-Q endorsement was mediated by ADRD neuropathologies and cognition (the Logical Memory II score). First, we examined the association of APOE e4 with the three critical pathologies, AD (represented by ADNPC), LBD, and CAA, in logistic regression models. Then, we examined the mediation models in the causal sequence as depicted in Figure 1: (Med1) APOE e4 as the independent variable, i.e., exposure, cognition as the outcome, and the three neuropathologies as the parallel multiple mediators; (Med2) each of the three neuropathologies served as the independent variables, cognition as the mediator, and each of the 12 NPIQ endorsements as the outcome controlling for E4; (Med3) APOE e4 as the independent variable, cognition as the mediator, and each NPI-Q endorsement as the outcome; (Med4) APOE e4 as the independent variable, each NPI endorsement as outcome, the three neuropathologies as parallel multiple mediators controlling for cognition.

Further, we conducted a more refined analysis using three AD pathology ratings (the AD ABC pathologies Thal stage, Braak stage and Neuritic plaque scores^18^ as multiple mediators, APOE e4 as the exposure, and Logical Memory II as the outcome. This was followed by a mediation analysis in which the A B C pathologies served as independent variables, Logical Memory II score as a mediator, and NPI items as the outcome measures and an analysis in which e4 was the exposure, the ABC pathologies were the mediators, and NPI items the outcomes. The ordinal ABC pathologies were treated as continuous variables for the purposes of these analysis.

We also explored the role of cognition by substituting Logical Memory II (verbal memory for stories after a delay) with MMSE (global cognitive status) in all mediations in a sensitivity analysis. We did this in order to determine if global cognitive or a more specific domain (memory) might be associated with specific mediation profiles.

For the estimation and statistical significance testing of the mediations, we used a model-based causal mediation analysis implemented in the mediation R package^22^. Each mediation analysis has two statistical models, the mediator model for the conditional distribution of the mediator given the exposure and covariates and the outcome model for the conditional distribution of the outcome given exposure, mediator, and covariates. These models are fitted separately, and then their fitted objects comprise the main inputs to the mediation function, which computes the estimated average causal mediation effect (ACME) and average direct effect (ADE). For binary NPIQ outcomes, the ACME and ADE are expressed as the increase in the probability that the subject endorses NPI symptom presence. For continuous outcomes such as cognition, the estimate effects are expressed as the increase or decrease in the outcomes in its unit. The 95% confidence intervals and p-values were computed based on bootstrapping with 5000 resamplings. Within each model in the causal sequence, paths were adjusted for significance using a False Discovery Rate (FDR) of p<.05.

For all analyses evaluating neuropathologies and NPI endorsements, we corrected for multiple comparisons with the Benjamini-Hochberg method, controlling for false discovery rate^23^. For post hoc and sensitivity analyses, uncorrected p values are reported.

We conducted a factor analysis of the NPI (Varimax orthogonal rotation) and identified three interpretable factors: psychosis, affective, frontal/drive. We used these descriptively to organize our reporting in Results and Discussion, but do not include them in mediation analyses.

## Results

### Cases

A total of 1199 cases with available NPIQ, cognition and neuropathological data were included. The flow chart of final case selection is in Supplementary Figure 1. The mean age at last visit was 80.3 years, and 687 participants (57%) were male. The mean years from final visit to death is 1.78. Nearly all ([95%]) individuals self-identified as White; data for other racial and ethnic groups are not reported owing to small numbers. 663 participants reported to be APOE e4 negative and 536 reported to be positive. These data and SDs are in Table 1, along with Logical Memory and MMSE scores. At the final clinic visit, the no known pathology group of 97 study participants was heterogenous with global Clinical Dementia Rating scores ranging from 1 to 3. The mean total NPIQ score was 5.45. For the three critical pathologies, the proportion having AD (70%) was the highest, followed by the presence of CAA (62%) and LBD (38%). The proportions of NPIQ symptom presence by E4 status are summarized in Table 2. Apathy was the most frequently reported symptom (45% of cases), while elation/euphoria the least (5%).

**Table 1.**
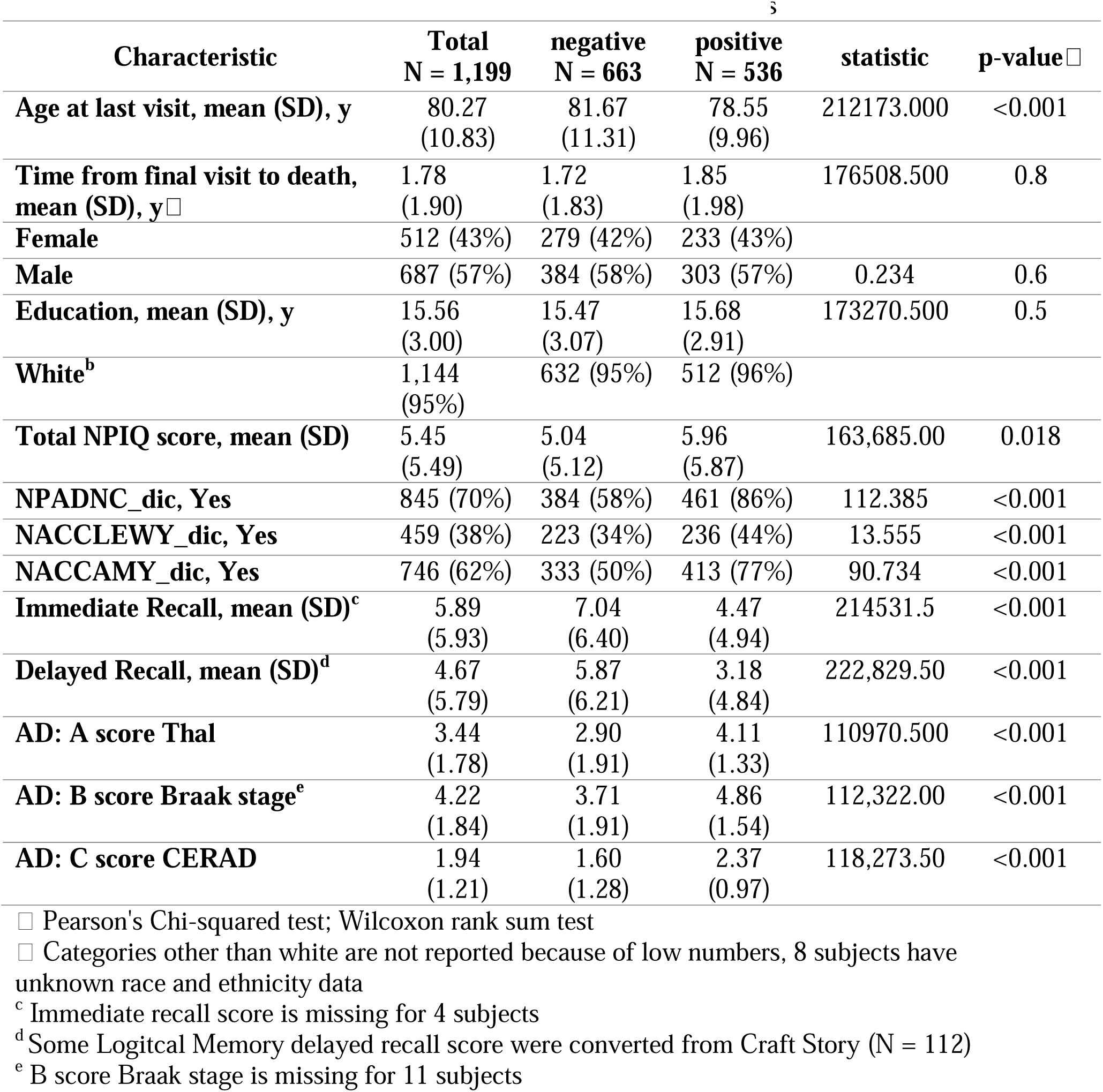
Demographic and Clinical Characteristics by APOE E4 Status.

| Characteristic | Total<br>N = 1,199 | negative<br>N = 663 | positive<br>N = 536 | statistic | p-value<sup>□</sup> |
| --- | --- | --- | --- | --- | --- |
| Age at last visit, mean (SD), y | 80.27<br>(10.83) | 81.67<br>(11.31) | 78.55<br>(9.96) | 212173.000 | <0.001 |
| Time from final visit to death, mean (SD), y<sup>□</sup> | 1.78<br>(1.90) | 1.72<br>(1.83) | 1.85<br>(1.98) | 176508.500 | 0.8 |
| Female | 512 (43%) | 279 (42%) | 233 (43%) |  |  |
| Male | 687 (57%) | 384 (58%) | 303 (57%) | 0.234 | 0.6 |
| Education, mean (SD), y | 15.56<br>(3.00) | 15.47<br>(3.07) | 15.68<br>(2.91) | 173270.500 | 0.5 |
| White<sup>b</sup> | 1,144<br>(95%) | 632 (95%) | 512 (96%) |  |  |
| Total NPIQ score, mean (SD) | 5.45<br>(5.49) | 5.04<br>(5.12) | 5.96<br>(5.87) | 163,685.00 | 0.018 |
| NPADNC_dic, Yes | 845 (70%) | 384 (58%) | 461 (86%) | 112.385 | <0.001 |
| NACCLEWY_dic, Yes | 459 (38%) | 223 (34%) | 236 (44%) | 13.555 | <0.001 |
| NACCAMY_dic, Yes | 746 (62%) | 333 (50%) | 413 (77%) | 90.734 | <0.001 |
| Immediate Recall, mean (SD)<sup>c</sup> | 5.89<br>(5.93) | 7.04<br>(6.40) | 4.47<br>(4.94) | 214531.5 | <0.001 |
| Delayed Recall, mean (SD)<sup>d</sup> | 4.67<br>(5.79) | 5.87<br>(6.21) | 3.18<br>(4.84) | 222,829.50 | <0.001 |
| AD: A score Thal | 3.44<br>(1.78) | 2.90<br>(1.91) | 4.11<br>(1.33) | 110970.500 | <0.001 |
| AD: B score Braak stage<sup>e</sup> | 4.22<br>(1.84) | 3.71<br>(1.91) | 4.86<br>(1.54) | 112,322.00 | <0.001 |
| AD: C score CERAD | 1.94<br>(1.21) | 1.60<br>(1.28) | 2.37<br>(0.97) | 118,273.50 | <0.001 |

□ Pearson's Chi-squared test; Wilcoxon rank sum test
□ Categories other than white are not reported because of low numbers, 8 subjects have unknown race and ethnicity data
<sup>c</sup> Immediate recall score is missing for 4 subjects
<sup>d</sup> Some Logitcal Memory delayed recall score were converted from Craft Story (N = 112)
<sup>e</sup> B score Braak stage is missing for 11 subjects

**Table 2.**
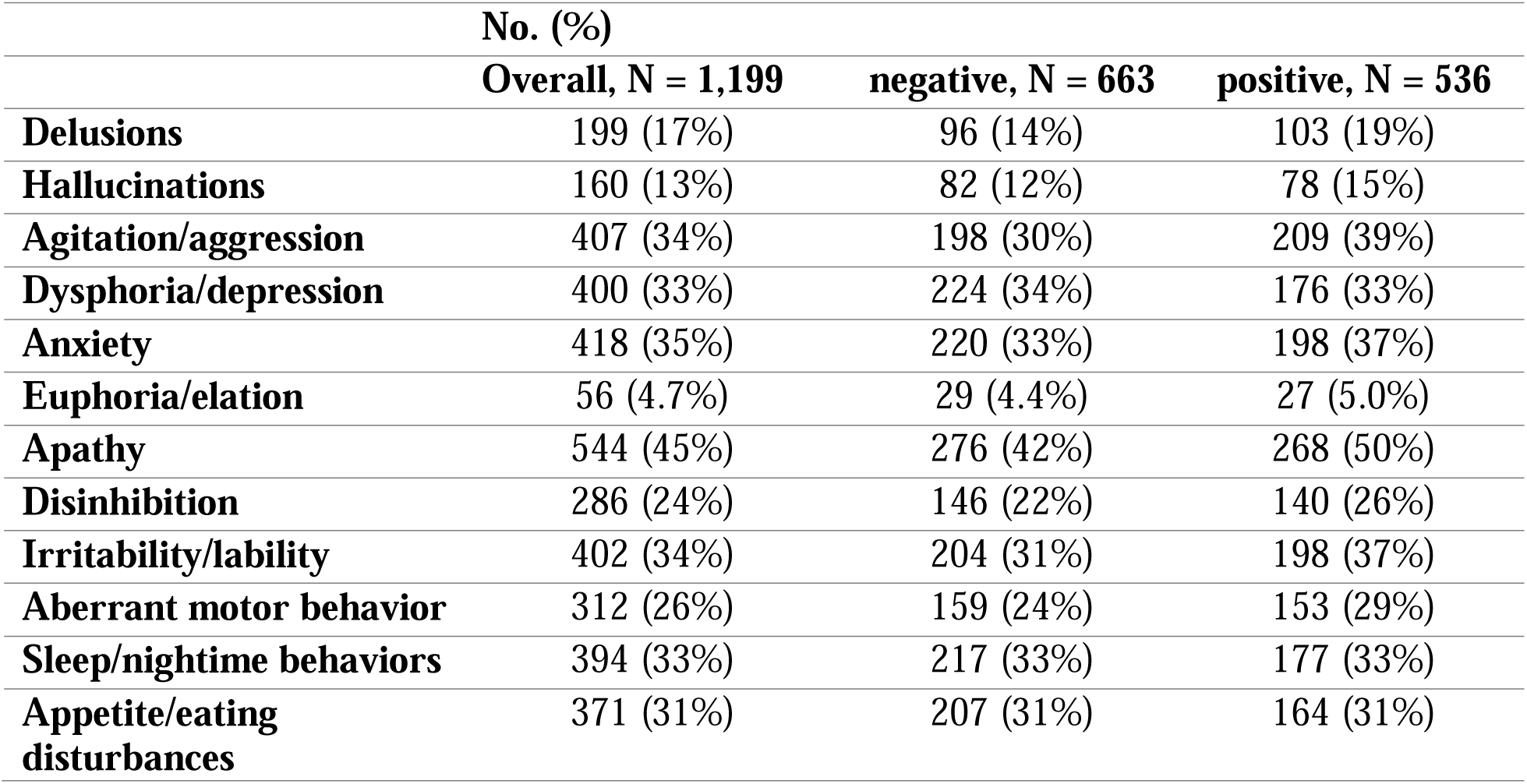
Frequencies of Neuropsychiatric Inventory Items (tabulated for presence) by APOE E4 Status.

### Preliminary Association Analyses

We first examined the association of APOE e4 with the three critical pathologies, AD (represented by ADNPC), LBD, and CAA, in multivariate logistic regression models. E4 was significantly associated with each pathology, controlled for all others, in keeping with the literature (Table 3). ADNPC demonstrated the largest association, followed by CAA and then LBD.

**Table 3.** Association of APOE E4 with ADRD pathologies. All logistic regression models adjusted for sex, age at visit, education and the time. OR=Odds Ratio.

| *Predictors* | *AD* |  |  | *LBD* |  |  | *CAA* |  |  |
| --- | --- | --- | --- | --- | --- | --- | --- | --- | --- |
|  | *OR* | *CI* | *p* | *OR* | *CI* | *p* | *OR* | *CI* | *p* |
| (Intercept) | 0.18 | 0.05 – 0.66 | 0.01 | 0.8 | 0.25 – 2.54 | 0.708 | 0.22 | 0.07 – 0.74 | 0.014 |
| Sex: Female | 1.27 | 0.96 – 1.68 | 0.1 | 0.79 | 0.61 – 1.01 | 0.06 | 1 | 0.77 – 1.30 | 0.979 |
| AGE at visit | 1.02 | 1.01 – 1.04 | <0.001 | 0.99 | 0.98 – 1.00 | 0.09 | 1.02 | 1.00 – 1.03 | 0.005 |
| Education(years) | 0.99 | 0.94 – 1.03 | 0.532 | 1.02 | 0.98 – 1.06 | 0.395 | 0.99 | 0.95 – 1.04 | 0.78 |
| last to death year | 1.17 | 1.08 – 1.28 | <0.001 | 1.1 | 1.04 – 1.18 | 0.002 | 1.14 | 1.07 – 1.23 | <0.001 |
| NACCAPOE 4: positive | 4.9 | 3.65 – 6.63 | <0.001 | 1.5 | 1.18 – 1.90 | 0.001 | 3.52 | 2.73 – 4.58 | <0.001 |

### Primary Mediation Analyses with Memory as a Mediator

In our primary mediation analyses, we examined Logical Memory II (delayed paragraph recall) serving as mediator.

#### Mediation 1

E4 was the independent variable, neuropathologies the mediators, and cognition (Logical Memory) the outcome. The direct effect of e4 was highly significant. Mediated effects were as follows: (AD: -1.075, 95% CI -1.42 to -0.76, p<0.001; LBD: -0.075 95% CI -0.168 to -0.009, p=0.025; CAA: -0.229, 95% CI -0.463 to -0.016, p=0.04), thus explaining 56% of the total effect all together. The relative magnitude of the mediators is shown in Figure 2 (slide 2). AD accounted for the largest portion (explaining 44% of the mediated effect)

**Figure 2.**
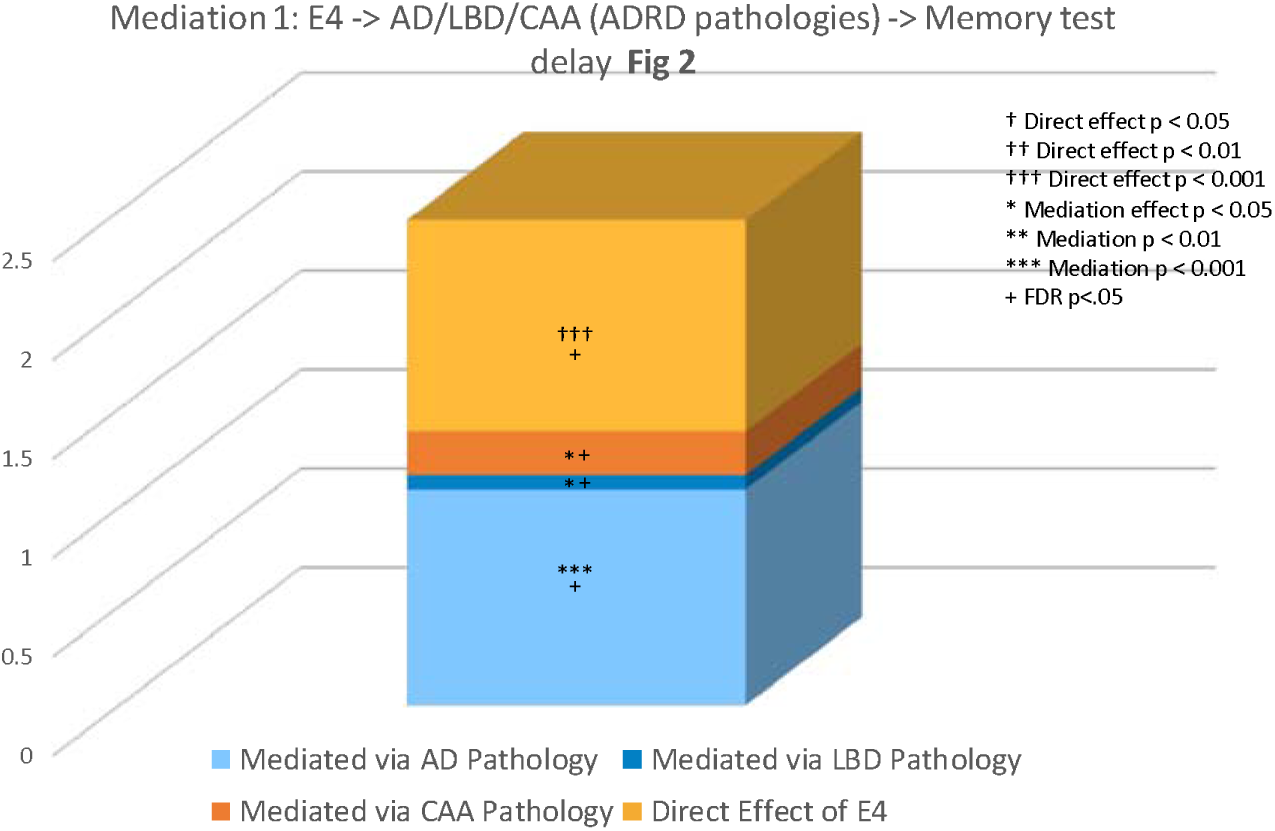
Mediation 1: E4 -> AD/LBD/CAA (ADRD pathologies) -> Memory test delay. The figure shows the respective magnitude of the indirect and direct path when APOE e4 served as the exposure, ADRD pathologies (AD, LB, and CAA (called AMY) the multiple mediators, and Logical Memory as the outcome. The thickness of the bars is proportional to the relative effect: AD pathology had the largest mediated effect on Logical Memory. Note also E4 had a significant direct effect. *=p<.5, **=p<.01, +=p<.05 FDR corrected

#### Mediation 2

We next examined the second mediation model in which the three pathologies served as the independent variables, Logical Memory delayed as the mediator, and NPI (all items) as the outcome (controlled for APOE). These are shown in Figures 3A-3C. The corresponding Table with CIs is in the Supplement (Table S1). For AD all items were significantly mediated by memory except for depression (Table S1). A single direct path to anxiety was present. For LBD all mediated paths were significant except depression. A single direct path to hallucinations was present. For CAA indirect paths were significant except depression, and all direct paths were not significant. Thus, there were widespread indirect paths from the pathologies (AD, LBD, CAA) to NPI items involving psychosis, affect, and frontal/drive symptoms.

**Figure 3A.**
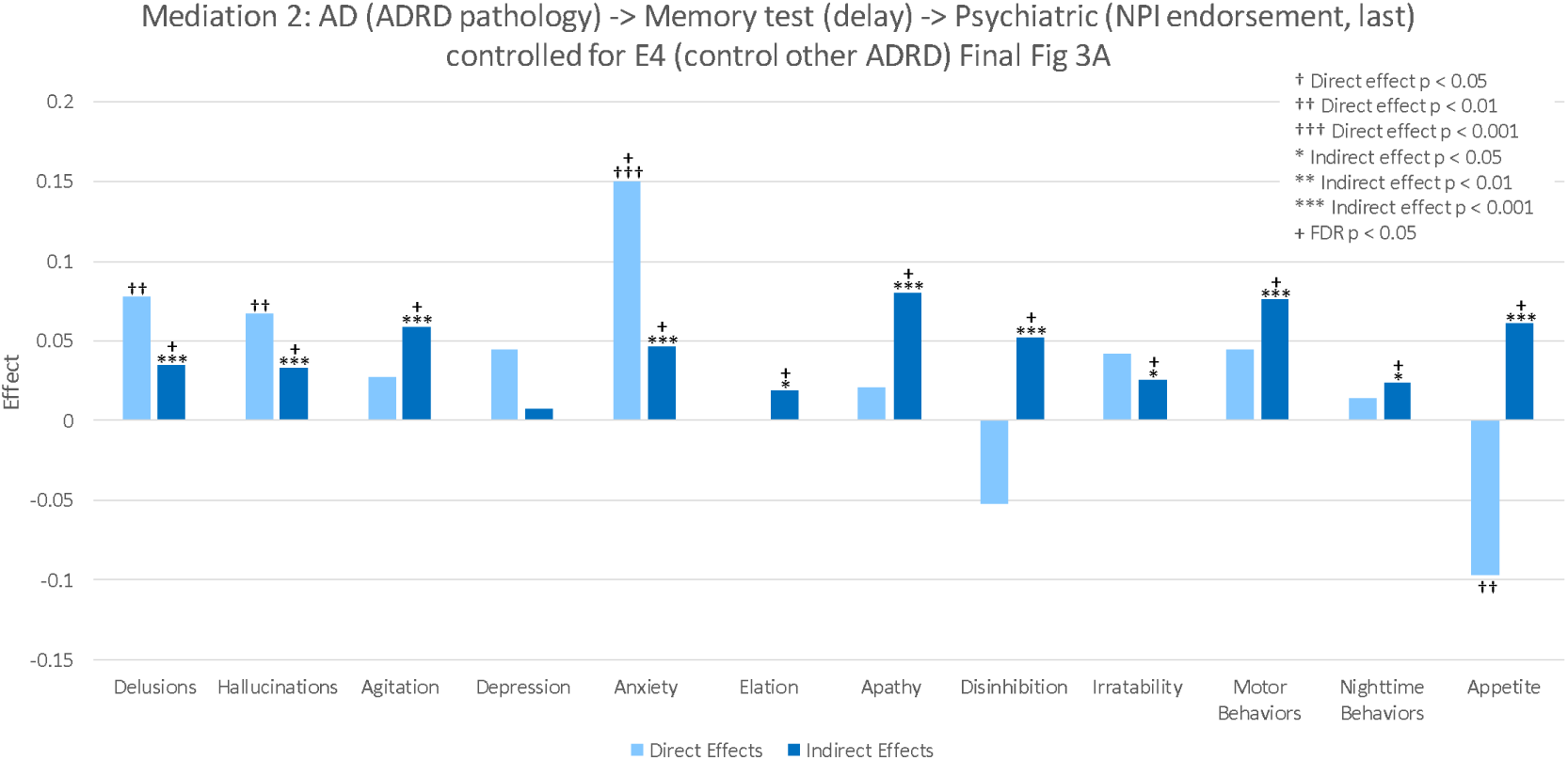
Mediation 2: AD (ADRD pathology) -> Memory test (delay) -> Psychiatric (NPI endorsement, last) controlled for E4 (control other ADRD) Mediations in which Logical Memory Delayed served as a mediator or outcome. Figures 3A 3B and 3C show the strength and significance level (FDR adjusted) of mediated and direct effects from the three ADRD neuropathologies to NPI items with memory as the mediator. 3A=AD, 3B=LBD, and 3C=CAA. Height of the bars represent the magnitude of the effect. Figure 3D shows the strength and significance of the direct effect from e4 (as the exposure) to each NPI items and the strength of the indirect paths in which memory served as the mediator. Figures 3E, 3F, and 3G show the strength and significance of the indirect effect between e4 (the exposure) and each items when the ADRD neuropathologies served as mediators (3E AD, 3F LBD, and 3F CAA) and the respective direct effects from e4 to NPI items.

**Figure 3B.**
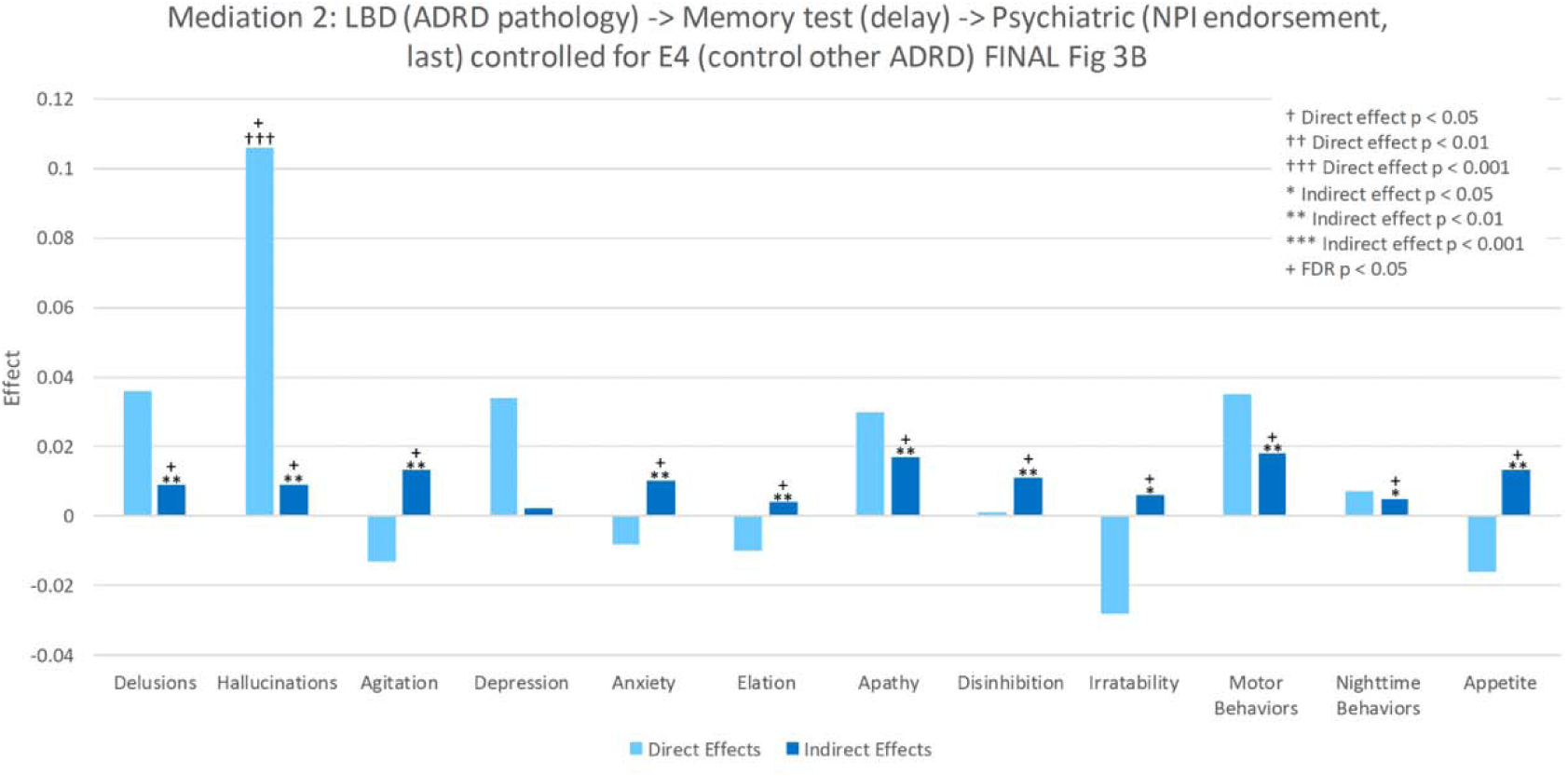
Mediation 2: LBD (ADRD pathology) -> Memory test (delay) -> Psychiatric (NPI endorsement, last) controlled for E4 (control other ADRD) Mediations in which Logical Memory Delayed served as a mediator or outcome. Figures 3A 3B and 3C show the strength and significance level (FDR adjusted) of mediated and direct effects from the three ADRD neuropathologies to NPI items with memory as the mediator. 3A=AD, 3B=LBD, and 3C=CAA. Height of the bars represent the magnitude of the effect. Figure 3D shows the strength and significance of the direct effect from e4 (as the exposure) to each NPI items and the strength of the indirect paths in which memory served as the mediator. Figures 3E, 3F, and 3G show the strength and significance of the indirect effect between e4 (the exposure) and each items when the ADRD neuropathologies served as mediators (3E AD, 3F LBD, and 3F CAA) and the respective direct effects from e4 to NPI items.

**Figure 3C.**
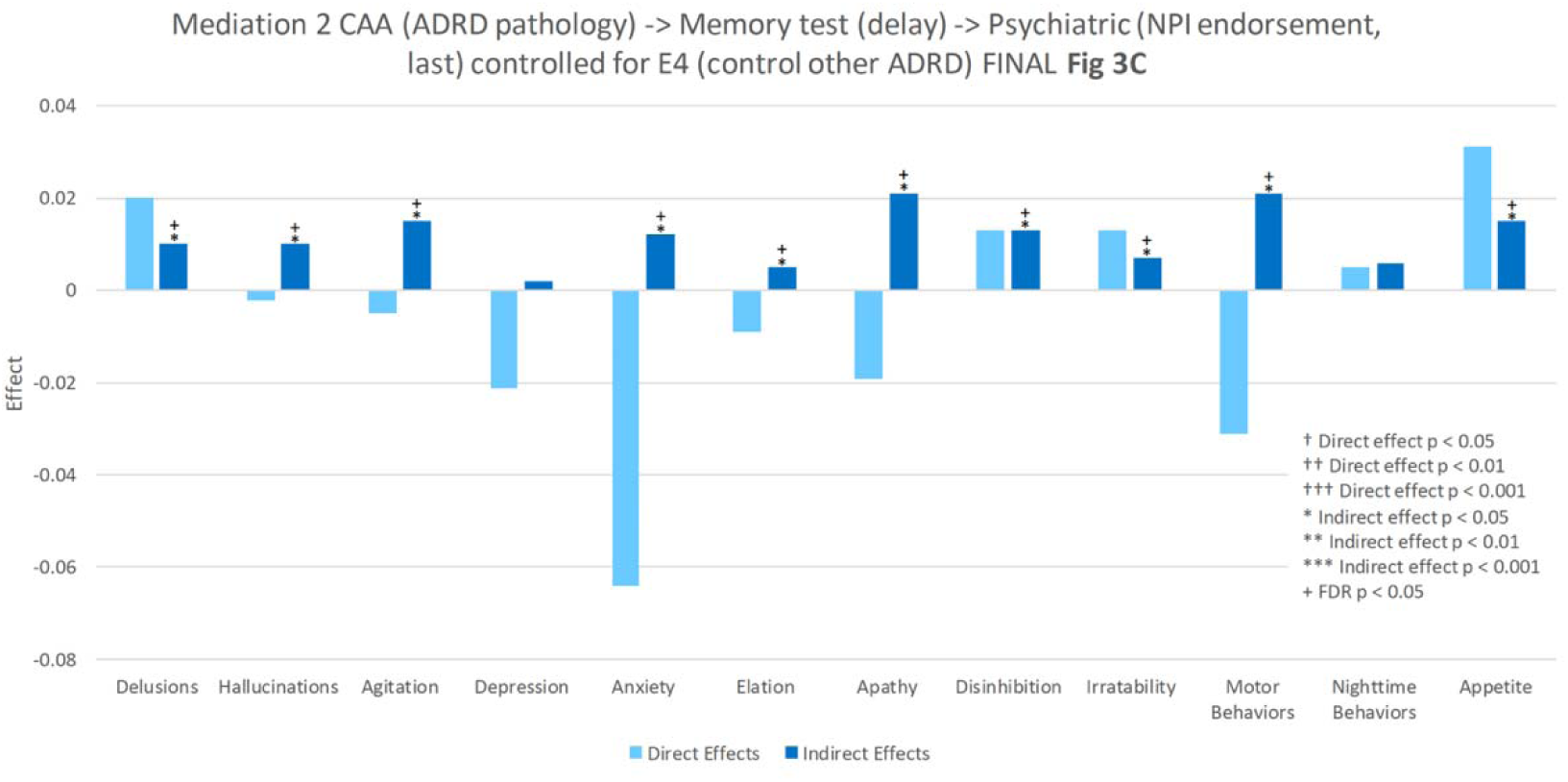
Mediation 2 CAA (ADRD pathology) -> Memory test (delay) -> Psychiatric (NPI endorsement, last) controlled for E4 (control other ADRD) Mediations in which Logical Memory Delayed served as a mediator or outcome. Figures 3A 3B and 3C show the strength and significance level (FDR adjusted) of mediated and direct effects from the three ADRD neuropathologies to NPI items with memory as the mediator. 3A=AD, 3B=LBD, and 3C=CAA. Height of the bars represent the magnitude of the effect. Figure 3D shows the strength and significance of the direct effect from e4 (as the exposure) to each NPI items and the strength of the indirect paths in which memory served as the mediator. Figures 3E, 3F, and 3G show the strength and significance of the indirect effect between e4 (the exposure) and each items when the ADRD neuropathologies served as mediators (3E AD, 3F LBD, and 3F CAA) and the respective direct effects from e4 to NPI items.

#### Mediation 3

The third model in this sequence included e4 as the independent variable, memory as the mediator, and NPI as the outcome controlling for all ADRD measures as shown in Fig 3D. CIs and total effects are in Supplementary Table S2. Memory mediated all associations of e4 with all NPI items except for depression. No direct path from e4 to NPI items was identified as significant.

**Figure 3D.**
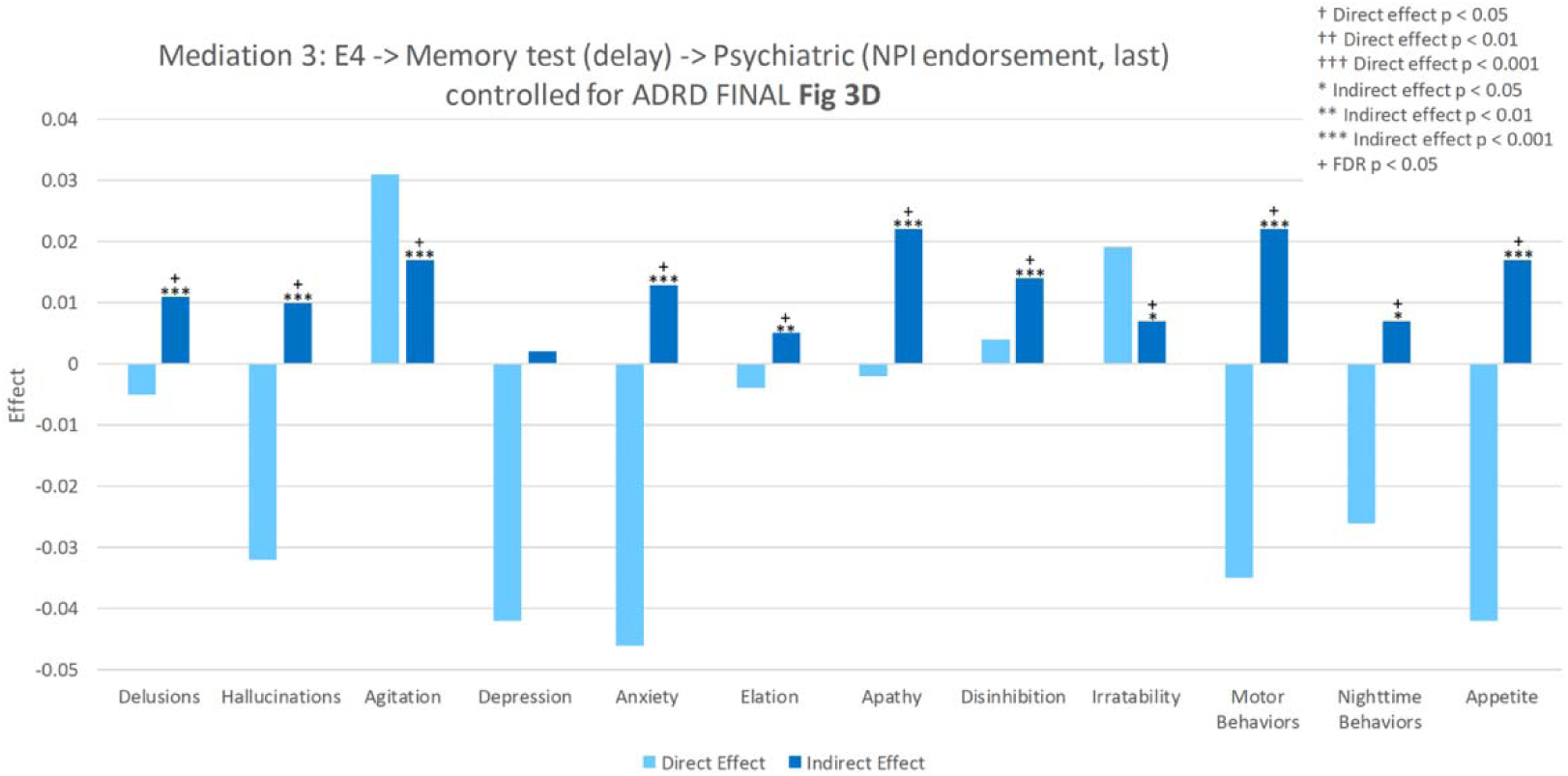
Mediation 3: E4 -> Memory test (delay) -> Psychiatric (NPI endorsement, last) controlled for ADRD. Mediations in which Logical Memory Delayed served as a mediator or outcome. Figures 3A 3B and 3C show the strength and significance level (FDR adjusted) of mediated and direct effects from the three ADRD neuropathologies to NPI items with memory as the mediator. 3A=AD, 3B=LBD, and 3C=CAA. Height of the bars represent the magnitude of the effect. Figure 3D shows the strength and significance of the direct effect from e4 (as the exposure) to each NPI items and the strength of the indirect paths in which memory served as the mediator. Figures 3E, 3F, and 3G show the strength and significance of the indirect effect between e4 (the exposure) and each items when the ADRD neuropathologies served as mediators (3E AD, 3F LBD, and 3F CAA) and the respective direct effects from e4 to NPI items.

#### Mediation 4

In the fourth mediation in which e4 served as the exposure, the three ADRD pathologies as mediators, and the NPI measures as outcomes, (controlled for memory) e4 had no direct effects. However, AD pathology had significant mediating effects on psychosis items, anxiety, and appetite. This is shown in Figure 3E, 3F, and 3G and tabled in Supplementary Table S3. The other pathologies did not demonstrate mediating effects in this model.

**Figure 3E.**
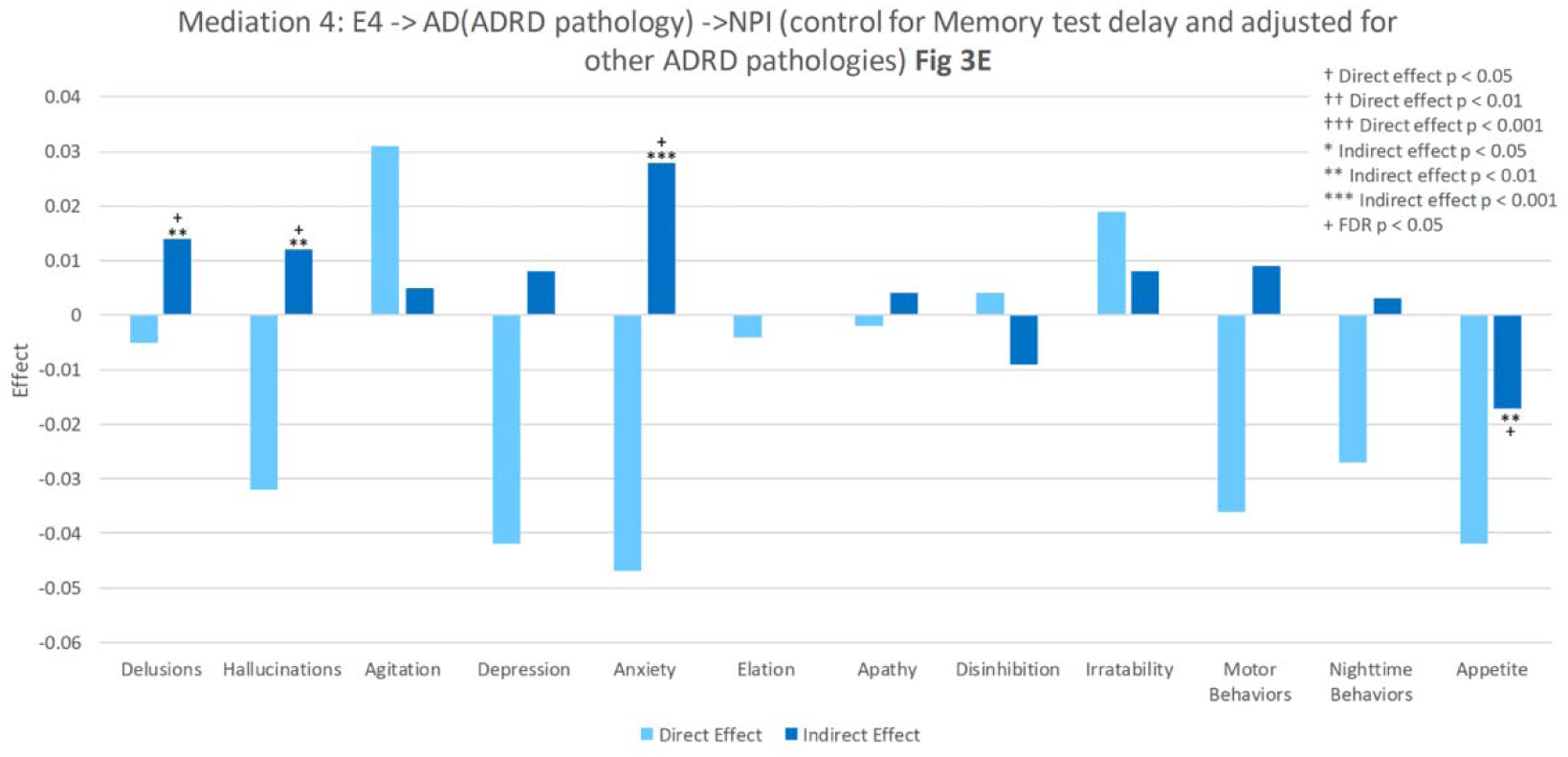
Mediation 4: E4 -> AD(ADRD pathology) ->NPI (control for Memory test delay and adjusted for other ADRD pathologies) Mediations in which Logical Memory Delayed served as a mediator or outcome. Figures 3A 3B and 3C show the strength and significance level (FDR adjusted) of mediated and direct effects from the three ADRD neuropathologies to NPI items with memory as the mediator. 3A=AD, 3B=LBD, and 3C=CAA. Height of the bars represent the magnitude of the effect. Figure 3D shows the strength and significance of the direct effect from e4 (as the exposure) to each NPI items and the strength of the indirect paths in which memory served as the mediator. Figures 3E, 3F, and 3G show the strength and significance of the indirect effect between e4 (the exposure) and each items when the ADRD neuropathologies served as mediators (3E AD, 3F LBD, and 3F CAA) and the respective direct effects from e4 to NPI items.

**Figure 3F.**
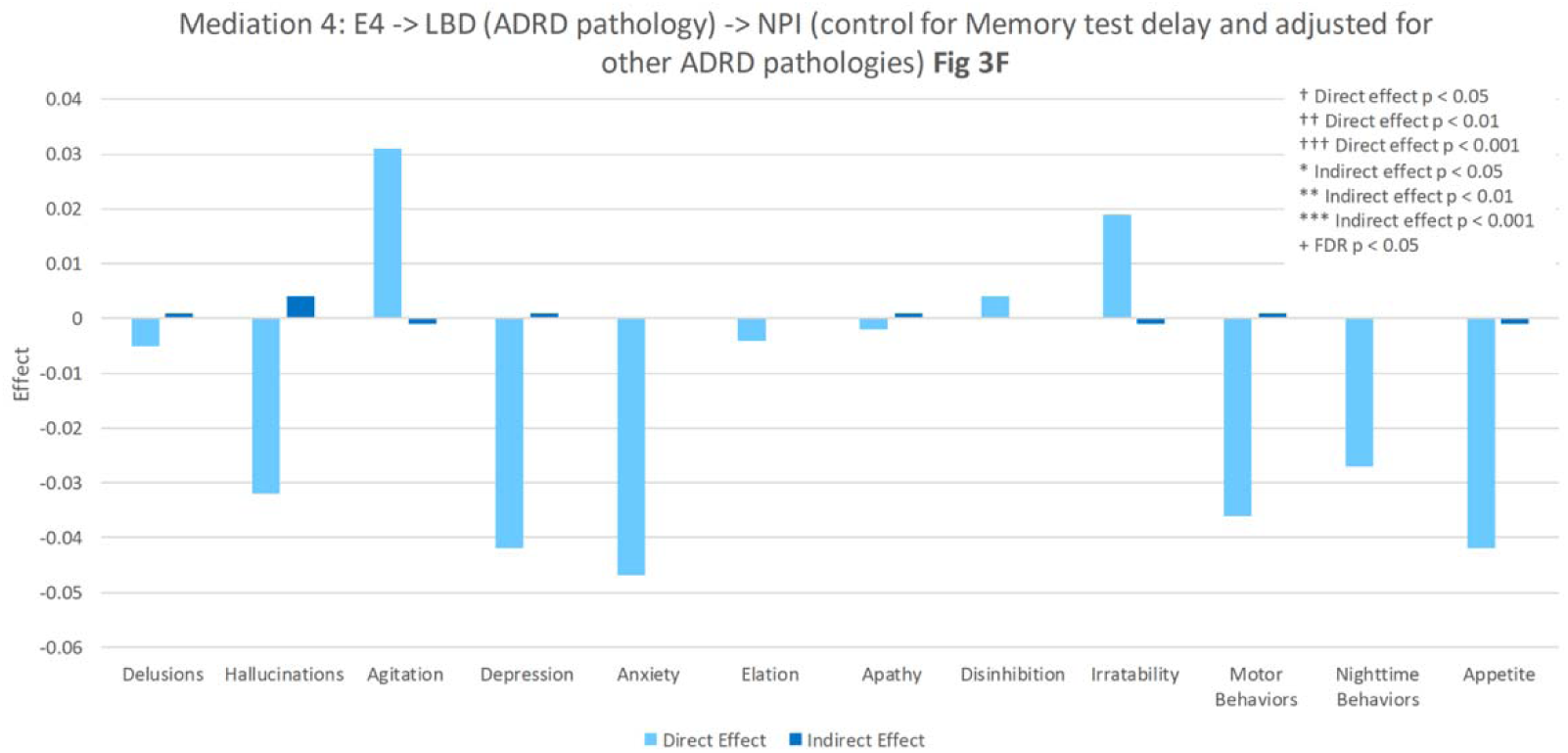
Mediation 4: E4 -> LBD (ADRD pathology) -> NPI (control for Memory test delay and adjusted for other ADRD pathologies) Mediations in which Logical Memory Delayed served as a mediator or outcome. Figures 3A 3B and 3C show the strength and significance level (FDR adjusted) of mediated and direct effects from the three ADRD neuropathologies to NPI items with memory as the mediator. 3A=AD, 3B=LBD, and 3C=CAA. Height of the bars represent the magnitude of the effect. Figure 3D shows the strength and significance of the direct effect from e4 (as the exposure) to each NPI items and the strength of the indirect paths in which memory served as the mediator. Figures 3E, 3F, and 3G show the strength and significance of the indirect effect between e4 (the exposure) and each items when the ADRD neuropathologies served as mediators (3E AD, 3F LBD, and 3F CAA) and the respective direct effects from e4 to NPI items.

**Figure 3G.**
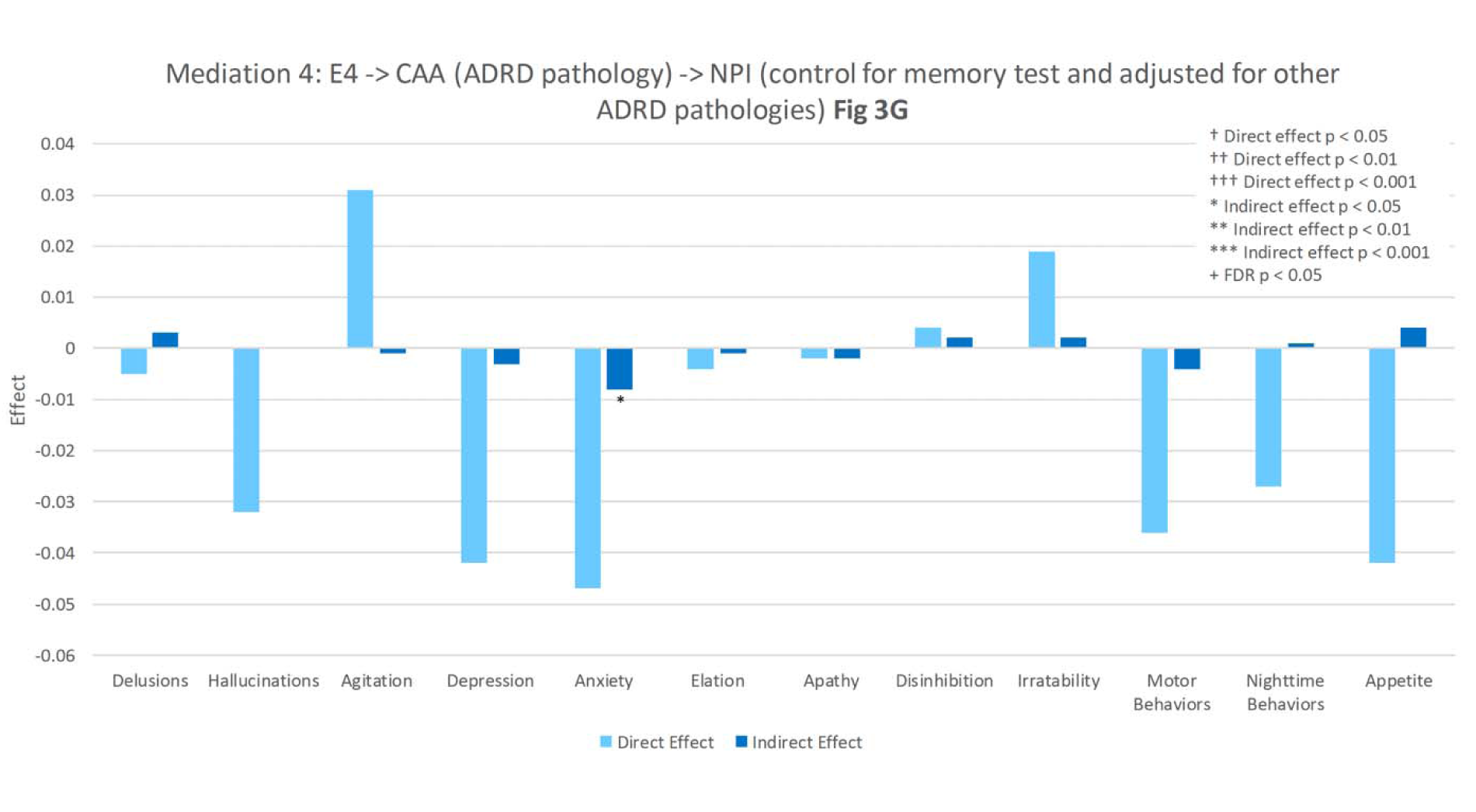
Mediation 4: E4 -> CAA (ADRD pathology) -> NPI (control for memory test and adjusted for other ADRD pathologies) Mediations in which Logical Memory Delayed served as a mediator or outcome. Figures 3A 3B and 3C show the strength and significance level (FDR adjusted) of mediated and direct effects from the three ADRD neuropathologies to NPI items with memory as the mediator. 3A=AD, 3B=LBD, and 3C=CAA. Height of the bars represent the magnitude of the effect. Figure 3D shows the strength and significance of the direct effect from e4 (as the exposure) to each NPI items and the strength of the indirect paths in which memory served as the mediator. Figures 3E, 3F, and 3G show the strength and significance of the indirect effect between e4 (the exposure) and each items when the ADRD neuropathologies served as mediators (3E AD, 3F LBD, and 3F CAA) and the respective direct effects from e4 to NPI items.

### Use of A B C Neuroathologies for Mediation

#### ABC Mediation 1

We conducted the next set of mediations in order to: 1. assess which of the AD pathologies (namely Thal plaque extent, Braak stage, or neuritic plaque density) were the main drivers of the AD mediation effect seen above in Med1-4, given its significance, and 2. assess if the above results were due to a threshold effect (given that in the above analyses pathologies were treated categorically as present/absent) or if AD A B C pathologies, treated as continuous variables, could similar significant results. Here AD A (Thal stage plaque extent), B (Braak tau tangle stage), and C (neuritic plaque density) AD pathology ratings served as mediators, e4 as the independent variable, and Logical Memory delayed score as the outcome. Braak stage (indirect effect=-1.225, 95% CI -1.662 to -0.824, p<0.001) and neuritic plaque density (indirect effect=-0.551, 95% CI -0.95 to-0.153, p=0.009) were significant mediators of e4 effects, with the former having the largest effect of the three pathologies (mediated proportion 0.49). Thal extent of diffuse plaques was not a significant mediator (indirect effect=0.102, 95% CI -0.284 to 0.505, p=0.61). A significant direct effect of e4 was present (direct effect=-0.83, 95% CI -1.41 to -0.24, p=0.009). The direct effect proportion of the total effect was 0.33 and significant at p<.01. Additionally, it was clear from these analyses that the mediated effect of neuropathology in the primary analyses was not simply due to a categorical approach as we also found it in this analysis when neuropathologies were treated as continuous values. This is reported in the Supplementary Table S4.

#### ABC Mediation 2

In the next analysis, A, B, and C served as the independent variables, Logical Memory II score as the mediator, and NPI items as the outcome variables. E4 was controlled. Braak stage and neuritic plaque score had significant associations when mediated by memory with all NPI items expect depression and irritability (Table 4). Significant coefficients ranged from .001 to .02. Interestingly, neuritic plaques had a direct effect on apathy and Braak stage had a direct effect on apathy and appetite (p,.05), but these were not significant at FDR p<.05. Amyloid plaque extent (Thal stage) was not associated with any NPI item either directly or indirectly.

**Table 4.**
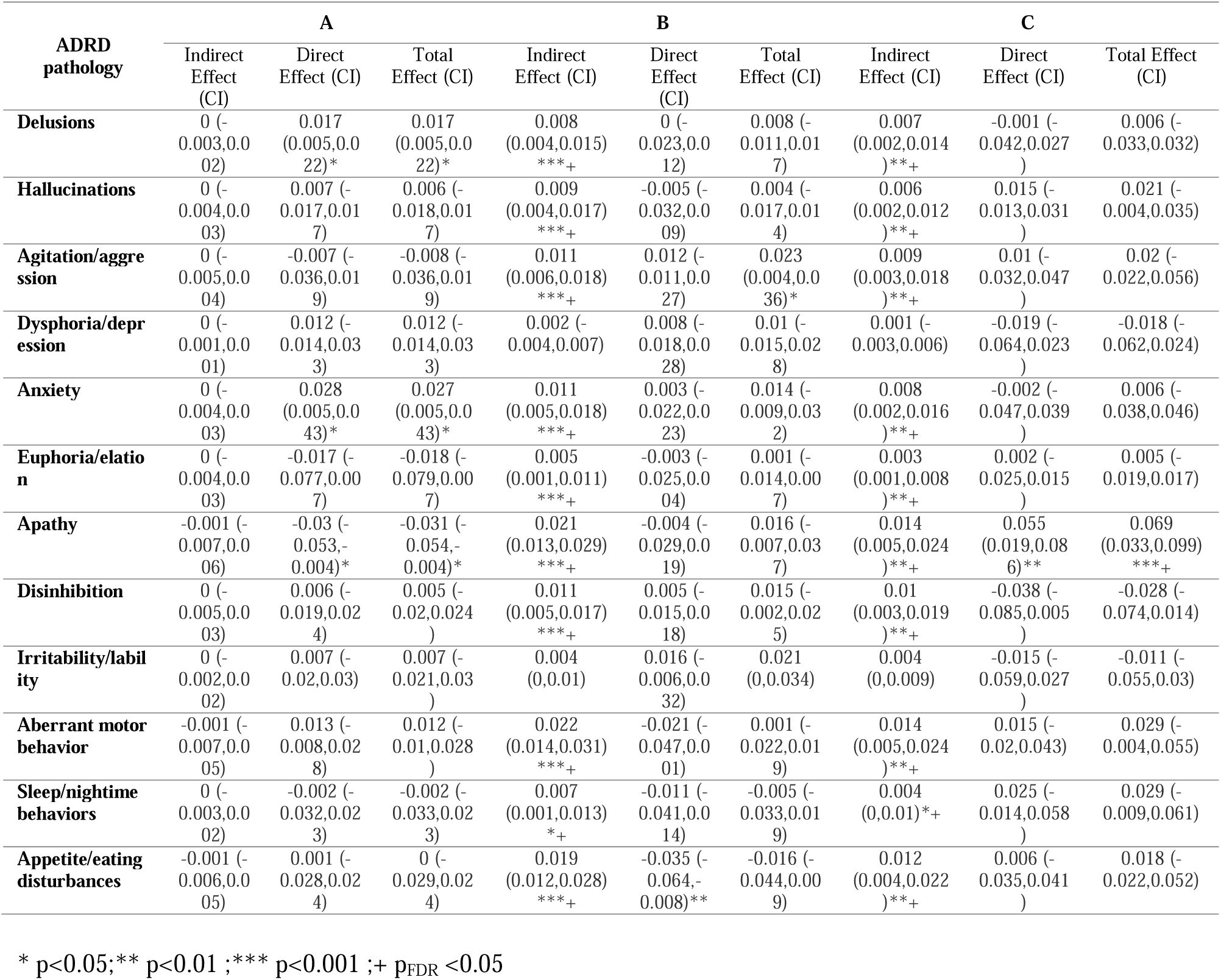
Mediation in which A (diffuse plaques), B (Braak stage), and C (neuritic plaque density) served as the independent variables, Logical Memory delayed score as the mediator, and NPI items as the outcome variables. E4 was controlled.

#### ABC Mediation 3

The third model in this sequence included e4 as the independent variable, Logical Memory II score as the mediator, and NPI as the outcome controlling for all ABC measures. All NPI item associations were significantly mediated by memory except depression and irritability. No direct path from e4 to NPI items was identified as significant, consistent with the earlier analyses above. See Supplement Table S5 with CIs.

#### ABC Mediation 4

E4 to ABC to NPI. The last model in this sequence included e4 as the independent variable, the ABC pathologies as the mediators, and NPI items as the outcome, memory controlled. No indirect paths were significant at FDR p<.05. In keeping with all other analyses, no direct path was present from e4 to any NPI item. See corresponding Table S6 with CIs.

### Sensitivity Analysis in which MMSE Replaced Logical Memory

We also conducted a parallel sensitivity analysis which examined the role of MMSE as a mediator (substituting as a measure of general cognitive status) when the three ADRD pathologies were involved, following the sequence of mediations in the Logical Memory analyses. In contradistinction to the Logical Memory analyses e4 was non-significant as having a direct effect on MMSE. Additionally, and strikingly, MMSE did not mediate NPI neurobehaviors when e4 was the exposure; this was in contradistinction to Logical Memory delayed. This may reflect the greater sensitivity of Logical Memory in detecting dementia as compared to the MMSE, given that memory is a cardinal feature of AD dementia. These results are described in the Supplement Parallel MMSE Sensitivity Analysis and reported in Supplementary Tables S7 and S8. Other models in this causal sequence are reported in Tables S9 and S10.

## Discussion

We derived three major findings from our analyses. While we found no evidence of direct effects of e4 on NPI items, all ADRD pathologies had direct effects on NPI items or effects on NPI mediated by cognition (either episodic memory for stories after a delay as tested on Logical Memory delayed or MMSE general cognitive status). This latter result suggests that many NPI behaviors in psychosis, affective or frontal/drive symptoms may be due to cognitive limitations and misinterpretations of the environment. It also suggests that therapeutic interventions that simplify and regularize the environment may be beneficial to patients with AD pathologies. In an important exception LBD had a direct effect on hallucinations in keeping with its known pathology in higher order visual areas in which functional compromises in BOLD or FDG PET and alpha synuclein aggregate pathology are present. Such pathology may directly result in tendencies toward pareidolias, incompletely formed hallucinations, and fully formed visual hallucinations^24, 25^. This finding also provides evidence in favor of the validity and sensitivity of our approach.

Second, when memory was the dependent measure, several striking differences emerged in both direct and indirect paths from e4 as compared to MMSE. E4 had a direct effect on memory, suggesting that the circuitry involved in episodic memory in the MTL including the hippocampus, may be susceptible to non-ADRD pathogenesis. No such direct effect was apparent when MMSE was the outcome. In particular and consistent with our results, it has been shown that e4 driven compromises of the BBB in hippocampal regions results in increasing memory impairments in individuals with mild cognitive impairment^26^. Nevertheless our work does not directly measure such hippocampal BBB breakdown and as such this discussion point remains speculative. In summary the MTL system may be susceptible to APOE e4 effects apart from amyloid and tau histopathology and may also be critical for the emergence of neurobehavioral abnormalities in psychosis, affect, and frontal drive domains. Additionally, memory mediated effects were generally more widespread than MMSE effects on NPI items, suggesting that a test assaying the relatively specific cognitive domain of episodic memory had a larger role in driving NPI items than a screening measures for general cognitive status and thus can act as a gateway to these maladaptive behaviors.

Third, in our analysis of the ADNC A B C pathologies, neuritic plaques and Braak stage (a measure of the progression of PHFs from medial temporal to neocortical areas) were the significant mediators of the association between e4 and memory, while Thal stage extent of diffuse plaques was not. When A B C pathologies were the independent variables and MMSE the mediator, again only neuritic plaques and Braak stage demonstrated significant indirect paths to multiple NPI items that included those involving psychosis, affect, and frontal/drive symptoms. Braak stage also had a direct effect on apathy, significant at p<.05, though it did not attain FDR significance. The result was consistent with an in vivo PET ligand study that revealed an association between tau orbitofrontal levels and apathy^27^.

It is thus of interest that we observed that not only tau Braak stage, but an amyloid pathology, had a significant effect on cognition. Our findings thus extend to neurobehavior a key earlier neuropathological observation in which neuritic plaques were more strongly associated with mental status than were diffuse plaques^28^. Speculatively our findings are consistent with recent in vitro work showed that neuritic plaques may disrupt synapses and result in large scale network failures^29^. However, we acknowledge that our findings do not directly measure network failure. Plaque extent using the Thal scale did not attain significance, suggesting that diffuse plaques were less “toxic” or otherwise less deleterious to neuronal function than were neuritic plaques.

In terms of methodological considerations, we elected to conduct a causal sequence of mediation analyses rather than a structural equation for the following reasons. 1. We did not hypothesize that a latent variable was present in our data. 2. Because both mediator and outcome variables were ordinal or binary, a mediation analysis is more stable than a structural equation model. 3. The sequence of mediation analyses that we conducted is transparent in that indirect mediation and direct paths are clearly specified and quantified at every stage.

Previously, we examined direct associations of ADNP, LBD, FTLD, and hippocampal sclerosis with NPI items^30^. Here we were interested in the association of APOE with NPI items and the potential mediating role of pathologies established as genetically associated with APOE e4 (AD, LB, CAA), and a potential mediating role of cognition (not examined in prior work), in a causal chain of analyses. In more refined analyses we also directly examined three AD histopathologies (amyloid plaque extent, neuritic plaques, Braak stage) as mediators; this was not explored in the prior study. Other findings from the NACC neuropathology data set suggested that psychotic symptoms in AD pathological cases were driven by Lewy body pathology^31–33^. Our work, using a different statistical technique involving formal mediation analyses in which we were also able to examine the independent contributions of AD ABC pathology and Lewy bodies, came to different conclusions.

In summary we designed an innovative approach for integrating genotypic, neuropathological, cognitive, and neurobehavioral data that yielded insights into neurodegenerative disorders. We found strong evidence for partial mediation of NPI symptoms by ADRD pathologies or by cognition. It was also observed that cognitive limitations that may have influenced understanding (or misunderstanding) the environment may be mediators of ADRD associations with NPI items. In addition, neuritic amyloid plaque levels and Braak stage were key in NPI associations when they were mediators or when they were the independent measure, suggesting the possibility that synaptic failure and larger scale network disruption play an important role in multiple neurobehavioral symptoms in dementia, including psychosis. Last, while we found little to support our hypothesis that e4 may have direct effects on cognition when we used MMSE as an outcome, such a relationship was found when Logical Memory delayed served as an outcome. This in turn suggests that medial temporal regions that support memory may be sensitive to non-amyloidogenic and non-tau related pathophysiological processes.

## Data Availability

All data produced in the present study are available upon reasonable request to the authors and approval from NACC consortium.

## Acknowledgments

TEG, SL, DPD, AT, SC, HK, and ER were supported by NIA grant R01AG051346.The authors report no conflicts of interest.

## Contributions

TEG conceived the study, interpreted data, and wrote the first draft. SL designed the statistical approach and conducted all statistical analyses and contributed to data interpretation. DD and HK made substantive scientific revisions in drafts of the ms. ZF conducted statistical analyses and provided technical support. AT, SC, and ER provided comments about data interpretation and provided technical support.

## Notes

### Competing Interest Statement

The authors have declared no competing interest.

