## supplementary tables and figures for "Effects of APOE e4 and Neuropathological Diagnoses on Neuropsychiatric Symptoms: Mediation Analyses and Likely Causation in an Integrated NACC Database"

**Supplementary Results and Discussion**

**Results**

**Primary Analyses: Logical Memory Delayed as a Mediator**

Table S1. Mediation 2 testing whether the effect of neuropathology on NPI is mediated by logical memory.

| ADRD pathology | AD | | | LEWY | | | CAA | | |
| --- | --- | --- | --- | --- | --- | --- | --- | --- | --- |
|  | Indirect Effect (CI) | Direct Effect (CI) | Total Effect (CI) | Indirect Effect (CI) | Direct Effect (CI) | Total Effect (CI) | Indirect Effect (CI) | Direct Effect (CI) | Total Effect (CI) |
| Delusions | 0.035 (0.019,0.054)***+ | 0.078 (0.027,0.126)** | 0.113 (0.065,0.16)***+ | 0.009 (0.002,0.017)**+ | 0.036 (-0.007,0.083) | 0.045 (0.002,0.092)* | 0.01 (0.002,0.02)*+ | 0.02 (-0.031,0.07) | 0.03 (-0.021,0.079) |
| Hallucinations | 0.033 (0.018,0.05)***+ | 0.067 (0.019,0.111)** | 0.099 (0.054,0.143)***+ | 0.009 (0.002,0.017)**+ | 0.106 (0.064,0.152)***+ | 0.115 (0.073,0.16)***+ | 0.01 (0.002,0.019)*+ | -0.002 (-0.053,0.047) | 0.007 (-0.043,0.056) |
| Agitation/aggression | 0.059 (0.036,0.084)***+ | 0.027 (-0.045,0.099) | 0.086 (0.018,0.155)* | 0.013 (0.004,0.024)**+ | -0.013 (-0.066,0.043) | 0 (-0.054,0.056) | 0.015 (0.004,0.03)*+ | -0.005 (-0.069,0.061) | 0.011 (-0.055,0.076) |
| Dysphoria/depression | 0.007 (-0.013,0.028) | 0.045 (-0.028,0.116) | 0.052 (-0.018,0.121) | 0.002 (-0.003,0.007) | 0.034 (-0.021,0.093) | 0.036 (-0.019,0.094) | 0.002 (-0.003,0.008) | -0.021 (-0.086,0.046) | -0.019 (-0.083,0.048) |
| Anxiety | 0.046 (0.026,0.068)***+ | 0.15 (0.083,0.216)***+ | 0.196 (0.133,0.256)***+ | 0.01 (0.003,0.02)**+ | -0.008 (-0.061,0.047) | 0.002 (-0.05,0.058) | 0.012 (0.003,0.025)*+ | -0.064 (-0.127,0.001) | -0.052 (-0.115,0.012) |
| Euphoria/elation | 0.019 (0.007,0.034)*+ | 0.001 (-0.038,0.035) | 0.02 (-0.014,0.052) | 0.004 (0.001,0.009)**+ | -0.01 (-0.038,0.019) | -0.006 (-0.033,0.023) | 0.005 (0.001,0.011)*+ | -0.009 (-0.047,0.025) | -0.004 (-0.041,0.03) |
| Apathy | 0.08 (0.055,0.108)***+ | 0.021 (-0.053,0.096) | 0.101 (0.03,0.173)*+ | 0.017 (0.005,0.031)**+ | 0.03 (-0.025,0.088) | 0.048 (-0.008,0.107) | 0.021 (0.005,0.038)*+ | -0.019 (-0.085,0.049) | 0.001 (-0.067,0.07) |
| Disinhibition | 0.052 (0.031,0.076)***+ | -0.052 (-0.117,0.011) | 0 (-0.06,0.06) | 0.011 (0.003,0.021)**+ | 0.001 (-0.047,0.052) | 0.012 (-0.037,0.063) | 0.013 (0.003,0.026)*+ | 0.013 (-0.044,0.07) | 0.026 (-0.031,0.083) |
| Irritability/lability | 0.026 (0.006,0.049)*+ | 0.042 (-0.033,0.115) | 0.068 (-0.002,0.137) | 0.006 (0.001,0.013)*+ | -0.028 (-0.082,0.03) | -0.023 (-0.077,0.035) | 0.007 (0.001,0.016)*+ | 0.013 (-0.052,0.08) | 0.02 (-0.045,0.085) |
| Aberrant motor behavior | 0.076 (0.052,0.102)***+ | 0.045 (-0.015,0.103) | 0.12 (0.063,0.176)***+ | 0.018 (0.005,0.031)**+ | 0.035 (-0.012,0.086) | 0.053 (0.005,0.105)* | 0.021 (0.005,0.038)*+ | -0.031 (-0.089,0.028) | -0.01 (-0.07,0.05) |
| Sleep/nightime behaviors | 0.024 (0.004,0.046)*+ | 0.014 (-0.06,0.087) | 0.038 (-0.033,0.107) | 0.005 (0,0.012)*+ | 0.007 (-0.049,0.066) | 0.012 (-0.043,0.071) | 0.006 (0,0.015)*+ | 0.005 (-0.062,0.074) | 0.011 (-0.056,0.079) |
| Appetite/eating disturbances | 0.061 (0.039,0.086)***+ | -0.097 (-0.17,-0.025)** | -0.036 (-0.107,0.033) | 0.013 (0.004,0.024)**+ | -0.016 (-0.068,0.039) | -0.003 (-0.056,0.053) | 0.015 (0.004,0.029)*+ | 0.031 (-0.032,0.095) | 0.046 (-0.018,0.109) |

* p<0.05 ;** p<0.01 ;*** p<0.001 ;+ p_FDR_ <0.05

Supplementary Table S2. Mediation 3 testing whether the effect of e4 on NPI is mediated by Logical Memory delayed controlling for neuropathology.

| NPI items | Indirect Effect (CI) | Direct Effect (CI) | Total Effect (CI) |
| --- | --- | --- | --- |
| Delusions | 0.011 (0.004,0.02)***+ | -0.005 (-0.048,0.039) | 0.006 (-0.037,0.05) |
| Hallucinations | 0.01 (0.004,0.019)***+ | -0.032 (-0.072,0.009) | -0.021 (-0.062,0.02) |
| Agitation/aggression | 0.017 (0.007,0.029)***+ | 0.031 (-0.024,0.087) | 0.047 (-0.007,0.105) |
| Dysphoria/depression | 0.002 (-0.003,0.008) | -0.042 (-0.098,0.015) | -0.04 (-0.096,0.018) |
| Anxiety | 0.013 (0.005,0.024)***+ | -0.046 (-0.099,0.01) | -0.033 (-0.086,0.023) |
| Euphoria/elation | 0.005 (0.002,0.011)**+ | -0.004 (-0.034,0.025) | 0.001 (-0.028,0.031) |
| Apathy | 0.022 (0.009,0.037)***+ | -0.002 (-0.057,0.055) | 0.02 (-0.037,0.078) |
| Disinhibition | 0.014 (0.006,0.025)***+ | 0.004 (-0.046,0.055) | 0.018 (-0.032,0.071) |
| Irritability/lability | 0.007 (0.001,0.016)*+ | 0.019 (-0.038,0.078) | 0.026 (-0.03,0.085) |
| Aberrant motor behavior | 0.022 (0.009,0.037)***+ | -0.035 (-0.082,0.014) | -0.013 (-0.062,0.037) |
| Sleep/nightime behaviors | 0.007 (0.001,0.014)*+ | -0.026 (-0.083,0.032) | -0.02 (-0.077,0.039) |
| Appetite/eating disturbances | 0.017 (0.007,0.028)***+ | -0.042 (-0.097,0.013) | -0.026 (-0.081,0.031) |

* p<0.05;** p<0.01 ;*** p<0.001 ;+ p_FDR_ <0.05

Supplementary Table S3. Mediation 4 analysis of e4 as the exposure, the three ADRD pathologies as mediators, and the NPIitems as outcomes (controlled for memory).

| ADRD pathology | AD | LEWY | CAA |  |  |
| --- | --- | --- | --- | --- | --- |
|  | Indirect Effect (CI) | Indirect Effect (CI) | Indirect Effect (CI) | Direct Effect (CI) | Total Effect |
| Delusions | 0.014 (0.004,0.024)**+ | 0.001 (-0.001,0.006) | 0.003 (-0.004,0.009) | -0.005 (-0.048,0.039) | 0.013 (-0.032,0.060) |
| Hallucinations | 0.012 (0.003,0.022)**+ | 0.004 (-0.003,0.013) | 0 (-0.007,0.006) | -0.032 (-0.073,0.009) | -0.016 (-0.059,0.027) |
| Agitation/aggression | 0.005 (-0.008,0.018) | -0.001 (-0.004,0.002) | -0.001 (-0.009,0.008) | 0.031 (-0.023,0.088) | 0.035 (-0.023,0.094) |
| Dysphoria/depression | 0.008 (-0.004,0.022) | 0.001 (-0.002,0.006) | -0.003 (-0.012,0.006) | -0.042 (-0.097,0.016) | -0.035 (-0.094,0.025) |
| Anxiety | 0.028 (0.014,0.044)***+ | 0 (-0.004,0.003) | -0.008 (-0.018,0)* | -0.047 (-0.1,0.01) | -0.027 (-0.084,0.032) |
| Euphoria/elation | 0 (-0.007,0.007) | 0 (-0.002,0.001) | -0.001 (-0.006,0.003) | -0.004 (-0.033,0.025) | -0.005 (-0.035,0.025) |
| Apathy | 0.004 (-0.009,0.017) | 0.001 (-0.002,0.006) | -0.002 (-0.012,0.006) | -0.002 (-0.057,0.056) | 0.000 (-0.058,0.061) |
| Disinhibition | -0.009 (-0.022,0.002) | 0 (-0.003,0.003) | 0.002 (-0.006,0.009) | 0.004 (-0.046,0.055) | -0.004 (-0.055,0.049) |
| Irritability/lability | 0.008 (-0.005,0.021) | -0.001 (-0.006,0.002) | 0.002 (-0.007,0.01) | 0.019 (-0.037,0.078) | 0.027 (-0.032,0.088) |
| Aberrant motor behavior | 0.009 (-0.002,0.021) | 0.001 (-0.001,0.006) | -0.004 (-0.013,0.003) | -0.036 (-0.085,0.015) | -0.030 (-0.082,0.023) |
| Sleep/nightime behaviors | 0.003 (-0.01,0.016) | 0 (-0.003,0.004) | 0.001 (-0.008,0.009) | -0.027 (-0.083,0.032) | -0.023 (-0.082,0.037) |
| Appetite/eating disturbances | -0.017 (-0.032,-0.005)**+ | -0.001 (-0.004,0.002) | 0.004 (-0.004,0.013) | -0.042 (-0.096,0.014) | -0.056 (-0.113,0.002) |

* p<0.05;** p<0.01 ;*** p<0.001 ;+ p_FDR_ <0.05

**Sensitivity Analyses in which AD A, B and C pathologies served as Mediators**

**Table S4. Mediation of AD ABC pathologies as mediators of the effect of APOE e4 on Logical Memory.**

| Indirect Effect (CI) | Diffuse plaques | 0.102 (-0.284, 0.505) |
| --- | --- | --- |
|  | **Braak stage** | -1.225 (-1.662,-0.824)*** |
|  | **Neuritic plaques** | -0.551 (-0.950, -0.153)** |
| Direct Effect (CI) | -0.826(-1.405,-0.239)** | |
| Total Effect (CI) | -2.500(-3.137,-1.877)*** | |

* p<0.05 ;** p<0.01 ;*** p<0.001

**Supplementary Table S5: Mediation of Logical Memory delayed on the effect of e4 on NPI items, controlled for ABC pathologies**

| NPI items | Indirect Effect (CI) | Direct Effect (CI) | Total Effect (CI) |
| --- | --- | --- | --- |
| Delusions | 0.008 (0.002,0.016)***+ | -0.01 (-0.053,0.033) | -0.002 (-0.045,0.042) |
| Hallucinations | 0.009 (0.002,0.017)***+ | -0.029 (-0.069,0.012) | -0.02 (-0.061,0.021) |
| Agitation/aggression | 0.011 (0.003,0.021)***+ | 0.018 (-0.036,0.075) | 0.029 (-0.025,0.086) |
| Dysphoria/depression | 0.001 (-0.003,0.007) | -0.047 (-0.102,0.011) | -0.046 (-0.101,0.013) |
| Anxiety | 0.009 (0.002,0.018)***+ | -0.06 (-0.112,-0.006)* | -0.051 (-0.104,0.005) |
| Euphoria/elation | 0.005 (0.001,0.01)***+ | -0.002 (-0.032,0.029) | 0.003 (-0.027,0.034) |
| Apathy | 0.016 (0.005,0.03)***+ | -0.002 (-0.058,0.055) | 0.014 (-0.043,0.072) |
| Disinhibition | 0.01 (0.003,0.02)***+ | 0 (-0.049,0.052) | 0.011 (-0.04,0.063) |
| Irritability/lability | 0.004 (0,0.01) | 0.011 (-0.045,0.069) | 0.015 (-0.041,0.074) |
| Aberrant motor behavior | 0.017 (0.005,0.031)***+ | -0.035 (-0.082,0.014) | -0.017 (-0.067,0.032) |
| Sleep/nightime behaviors | 0.005 (0,0.012)***+ | -0.03 (-0.086,0.028) | -0.025 (-0.081,0.034) |
| Appetite/eating disturbances | 0.014 (0.004,0.026)***+ | -0.037 (-0.09,0.018) | -0.023 (-0.078,0.033) |

**Supplementary Table S6: Mediation by ABC scores on the effect of E4 on NPI items, controlled for Logical Memory**

| ADRD pathology |  | Indirect Effect (CI) |  |  |  |
| --- | --- | --- | --- | --- | --- |
|  | **Diffuse Plaques** | **Braak** | **Neuritic plaques** | **Direct Effect (CI)** | **Total Effect (CI)** |
| Delusions | 0.008 (0.002,0.017)** | 0 (-0.004,0.005) | 0 (-0.002,0.003) | -0.01 (-0.053,0.034) | -0.001 (-0.047,0.044) |
| Hallucinations | 0.003 (-0.004,0.01) | -0.001 (-0.005,0.004) | 0.001 (-0.001,0.005) | -0.029 (-0.07,0.012) | -0.026 (-0.068,0.017) |
| Agitation/aggression | -0.002 (-0.009,0.006) | 0.003 (-0.002,0.009) | 0.001 (-0.002,0.004) | 0.018 (-0.036,0.076) | 0.02 (-0.037,0.078) |
| Dysphoria/depression | 0.004 (-0.003,0.012) | 0.002 (-0.003,0.008) | -0.001 (-0.004,0.001) | -0.047 (-0.102,0.012) | -0.042 (-0.1,0.017) |
| Anxiety | 0.009 (0.002,0.018)* | 0.001 (-0.004,0.006) | 0 (-0.003,0.003) | -0.061 (-0.114,-0.005)* | -0.051 (-0.107,0.006) |
| Euphoria/elation | -0.002 (-0.007,0.003) | 0 (-0.003,0.002) | 0 (-0.001,0.002) | -0.002 (-0.032,0.029) | -0.004 (-0.035,0.027) |
| Apathy | -0.008 (-0.017,-0.001)* | -0.001 (-0.006,0.004) | 0.003 (-0.002,0.009) | -0.002 (-0.058,0.056) | -0.009 (-0.066,0.05) |
| Disinhibition | 0.002 (-0.004,0.009) | 0.001 (-0.003,0.006) | -0.001 (-0.006,0.001) | 0 (-0.05,0.053) | 0.002 (-0.05,0.055) |
| Irritability/lability | 0.002 (-0.005,0.01) | 0.004 (-0.001,0.01) | -0.001 (-0.004,0.002) | 0.011 (-0.045,0.07) | 0.017 (-0.041,0.076) |
| Aberrant motor behavior | 0.004 (-0.002,0.012) | -0.004 (-0.009,0) | 0.001 (-0.001,0.004) | -0.036 (-0.086,0.015) | -0.035 (-0.086,0.016) |
| Sleep/nightime behaviors | 0 (-0.008,0.007) | -0.002 (-0.007,0.003) | 0.001 (-0.001,0.005) | -0.03 (-0.087,0.029) | -0.031 (-0.089,0.028) |
| Appetite/eating disturbances | 0 (-0.007,0.008) | -0.006 (-0.012,-0.001)* | 0 (-0.002,0.003) | -0.037 (-0.091,0.019) | -0.042 (-0.097,0.014) |

**Sensitivity Analyses in which MMSE Served as a Mediator, Replacing logical Memory Delayed**

**Supplementary Table S7. Mediation analysis of e4 as the exposure, ADRD pathologies as mediators, and MMSE as the outcome.**

| Indirect Effect (CI) | AD | -1.454 (-1.860,-1.081)***+ |
| --- | --- | --- |
|  | **LEWY** | -0.122 (-0.248,-0.028)**+ |
|  | **AMY** | -0.406 (-0.716,-0.117)**+ |
| Direct Effect (CI) | -0.235 (-1.048,0.579) | |
| Total Effect (CI) | -2.217 (-3.068,-1.363)***+ | |

**Supplementary Table S8. Mediation analysis of e4 as the exposure, MMSE pathologies as a mediator and NPI as the outcomes (controlling for ADRD pathologies)**

| NPI items | Indirect Effect (CI) | Direct Effect (CI) | Total Effect (CI) |
| --- | --- | --- | --- |
| Delusions | 0.001 (-0.002,0.004) | 0.006 (-0.033,0.047) | 0.007 (-0.032,0.048) |
| Hallucinations | 0.001 (-0.002,0.005) | -0.008 (-0.046,0.03) | -0.007 (-0.045,0.031) |
| Agitation/aggression | 0.002 (-0.004,0.008) | 0.045 (-0.006,0.098) | 0.047 (-0.004,0.1) |
| Dysphoria/depression | -0.001 (-0.004,0.002) | -0.032 (-0.083,0.02) | -0.033 (-0.084,0.02) |
| Anxiety | 0.001 (-0.002,0.005) | -0.024 (-0.075,0.028) | -0.024 (-0.074,0.028) |
| Euphoria/elation | 0.001 (-0.001,0.003) | -0.008 (-0.036,0.02) | -0.008 (-0.036,0.021) |
| Apathy | 0.003 (-0.006,0.012) | 0.017 (-0.035,0.072) | 0.02 (-0.033,0.075) |
| Disinhibition | 0.001 (-0.003,0.006) | 0.013 (-0.034,0.061) | 0.014 (-0.033,0.063) |
| Irritability/lability | 0.001 (-0.002,0.004) | 0.023 (-0.029,0.076) | 0.023 (-0.028,0.076) |
| Aberrant motor behavior | 0.002 (-0.006,0.011) | 0.002 (-0.046,0.051) | 0.004 (-0.044,0.054) |
| Sleep/nightime behaviors | 0.001 (-0.002,0.005) | -0.018 (-0.07,0.036) | -0.017 (-0.069,0.037) |
| Appetite/eating disturbances | 0.001 (-0.002,0.004) | -0.024 (-0.075,0.028) | -0.024 (-0.075,0.029) |

Supplement sensitivity analysis Med 1. We examined the second mediation model in which the three pathologies served as the independent variables, cognition as the mediator, and NPI (all items) as the outcomes controlling for APOE e4. For ADNPC effects on multiple NPI items were significantly mediated by MMSE (Supplementary Table S9). These included psychosis, affective and frontal type behaviors. The significant indirect effects ranged from .01 to .05, indicating that when neuropathology was present, the probability of having NPI symptoms increased by 1%-5% when mediated by the MMSE. In particular, the ADNC increased the probability of apathy by 5% through cognition. Direct paths were significant for delusions, hallucinations, and anxiety.

For LBD multiple indirect effects were significant, but values were smaller than for ADNC; they ranged from less than .01 to .01. A single direct path was present from LBD to hallucinations.

For CAA, multiple significant indirect paths were present across psychosis, affective, and frontal neurobehavioral, with coefficients ranging from less than .01 to .015. No direct effects were present. There were multiple instances of inconsistent mediation in which direct effects were negative and indirect effects positive, suggesting the presence of suppressor effects. This is shown in supplementary Table S9.

**Supplementary Table S9: Mediation analysis in which AD/LBD/CAA (ADRD pathology) served as the exposures, MMSE as the mediator, and NPI items as the outcome, controlled for E4**

| ADRD pathology | AD | | | LBD | | | CAA | | |
| --- | --- | --- | --- | --- | --- | --- | --- | --- | --- |
|  | **Indirect Effect (CI)** | **Direct Effect (CI)** | **Total Effect (CI)** | **Indirect Effect (CI)** | **Direct Effect (CI)** | **Total Effect (CI)** | **Indirect Effect (CI)** | **Direct Effect (CI)** | **Total Effect (CI)** |
| Delusions | 0.015 (0.005,0.026)**+ | 0.098 (0.051,0.14)***+ | 0.112 (0.07,0.153)***+ | 0.004 (0.001,0.009)**+ | 0.034 (-0.006,0.077) | 0.038 (-0.002,0.081) | 0.005 (0.001,0.01)**+ | 0.021 (-0.025,0.067) | 0.026 (-0.02,0.071) |
| Hallucinations | 0.018 (0.009,0.029)***+ | 0.071 (0.026,0.113)**+ | 0.089 (0.048,0.13)***+ | 0.005 (0.001,0.01)**+ | 0.089 (0.05,0.13)***+ | 0.094 (0.055,0.136)***+ | 0.006 (0.002,0.012)**+ | -0.01 (-0.057,0.036) | -0.004 (-0.05,0.041) |
| Agitation/aggression | 0.034 (0.019,0.051)***+ | 0.062 (-0.005,0.128) | 0.096 (0.033,0.158)**+ | 0.008 (0.002,0.015)**+ | -0.018 (-0.068,0.034) | -0.01 (-0.06,0.042) | 0.01 (0.003,0.019)**+ | -0.003 (-0.063,0.059) | 0.007 (-0.053,0.068) |
| Dysphoria/depression | -0.014 (-0.03,0.001) | 0.046 (-0.021,0.111) | 0.032 (-0.033,0.096) | -0.003 (-0.008,0) | 0.033 (-0.017,0.086) | 0.03 (-0.02,0.083) | -0.004 (-0.01,0) | -0.001 (-0.06,0.061) | -0.005 (-0.064,0.057) |
| Anxiety | 0.016 (0.002,0.031)*+ | 0.158 (0.095,0.219)***+ | 0.174 (0.114,0.231)***+ | 0.004 (0,0.009)*+ | -0.029 (-0.078,0.023) | -0.025 (-0.074,0.027) | 0.005 (0,0.012)*+ | -0.042 (-0.101,0.02) | -0.037 (-0.095,0.024) |
| Euphoria/elation | 0.009 (0.002,0.018)*+ | 0.011 (-0.026,0.044) | 0.02 (-0.014,0.053) | 0.002 (0,0.005)*+ | -0.015 (-0.042,0.013) | -0.013 (-0.039,0.016) | 0.003 (0,0.007)*+ | -0.013 (-0.052,0.024) | -0.01 (-0.049,0.027) |
| Apathy | 0.052 (0.035,0.072)***+ | -0.005 (-0.072,0.064) | 0.047 (-0.018,0.114) | 0.012 (0.004,0.022)**+ | 0.033 (-0.019,0.087) | 0.045 (-0.007,0.1) | 0.015 (0.005,0.027)**+ | -0.028 (-0.089,0.036) | -0.012 (-0.075,0.05) |
| Disinhibition | 0.024 (0.011,0.039)***+ | -0.016 (-0.076,0.043) | 0.008 (-0.049,0.065) | 0.006 (0.001,0.012)**+ | -0.009 (-0.055,0.04) | -0.004 (-0.049,0.045) | 0.007 (0.002,0.014)**+ | 0.001 (-0.054,0.057) | 0.008 (-0.046,0.063) |
| Irritability/lability | 0.011 (-0.004,0.027) | 0.049 (-0.018,0.116) | 0.06 (-0.004,0.124) | 0.003 (-0.001,0.007) | -0.028 (-0.077,0.025) | -0.025 (-0.075,0.028) | 0.003 (-0.001,0.009) | 0.026 (-0.033,0.088) | 0.029 (-0.03,0.09) |
| Aberrant motor behavior | 0.047 (0.032,0.064)***+ | 0.046 (-0.014,0.105) | 0.093 (0.036,0.148)**+ | 0.011 (0.004,0.021)**+ | 0.028 (-0.019,0.078) | 0.04 (-0.007,0.089) | 0.014 (0.004,0.025)**+ | -0.026 (-0.083,0.033) | -0.012 (-0.07,0.046) |
| Sleep/nightime behaviors | 0.017 (0.002,0.034)*+ | 0.017 (-0.05,0.084) | 0.035 (-0.031,0.098) | 0.004 (0,0.01)*+ | 0.015 (-0.036,0.069) | 0.019 (-0.032,0.073) | 0.005 (0,0.012)*+ | 0.004 (-0.057,0.067) | 0.009 (-0.052,0.071) |
| Appetite/eating disturbances | 0.012 (-0.003,0.027) | -0.045 (-0.112,0.023) | -0.033 (-0.099,0.032) | 0.003 (-0.001,0.007) | 0 (-0.05,0.052) | 0.002 (-0.047,0.055) | 0.003 (-0.001,0.009) | 0.025 (-0.034,0.085) | 0.028 (-0.031,0.087) |

Supplement sensitivity analysis Med2. Last, we examined the mediation model in which e4 served as the independent variable and neuropathologies as the mediators with NPI items as the outcome controlling for MMSE. AD pathology mediated hallucinations, delusions, affective symptoms and anxiety with indirect path weights ranging from about .02 to .05, indicating that e4 exposures increased the probability of having NPI symptoms by 3-5% through ADNC (Table S10). No direct path was significant. When LBD was a mediator, only hallucinations were mediated significantly in the presence of an e4 background. No significant direct paths were present. For CAA no indirect nor direct paths were significant.

**Supplementary Table S10: Mediation analysis in which E4 Served as the Exposure, AD/LBD/CAA (ADRD pathologies) as the Mediators, and NPI items as the Outcomes (controlled for MMSE)**

| ADRD pathology |  | Indirect Effect (CI) |  | Direct Effect (CI) | Total Effect (CI) |
| --- | --- | --- | --- | --- | --- |
|  | **AD** | **LEWY** | **CAA** |  |  |
| Delusions | 0.03 (0.016,0.044)***+ | 0.004 (-0.001,0.009) | 0.006 (-0.008,0.019) | 0.006 (-0.032,0.047) | 0.046 (-0.001,0.095) |
| Hallucinations | 0.023 (0.008,0.037)**+ | 0.01 (0.004,0.017)**+ | -0.003 (-0.017,0.01) | -0.008 (-0.045,0.03) | 0.021 (-0.024,0.067) |
| Agitation/aggression | 0.019 (-0.001,0.04) | -0.002 (-0.008,0.004) | -0.001 (-0.019,0.016) | 0.044 (-0.005,0.097) | 0.061 (-0.003,0.123) |
| Dysphoria/depression | 0.014 (-0.006,0.033) | 0.004 (-0.002,0.01) | 0 (-0.018,0.017) | -0.033 (-0.083,0.019) | -0.016 (-0.078,0.046) |
| Anxiety | 0.048 (0.028,0.069)***+ | -0.003 (-0.01,0.002) | -0.012 (-0.03,0.005) | -0.025 (-0.074,0.027) | 0.008 -(0.053,0.069) |
| Euphoria/elation | 0.003 (-0.008,0.014) | -0.002 (-0.005,0.001) | -0.004 (-0.016,0.007) | -0.008 (-0.037,0.021) | -0.010 (-0.047,0.024) |
| Apathy | -0.001 (-0.021,0.019) | 0.004 (-0.002,0.01) | -0.008 (-0.026,0.009) | 0.017 (-0.034,0.071) | 0.011 (-0.053,0.075) |
| Disinhibition | -0.005 (-0.024,0.013) | -0.001 (-0.007,0.004) | 0 (-0.016,0.016) | 0.013 (-0.034,0.061) | 0.007 (-0.050,0.064) |
| Irritability/lability | 0.015 (-0.005,0.035) | -0.003 (-0.01,0.002) | 0.008 (-0.01,0.025) | 0.022 (-0.028,0.075) | 0.042 (-0.021,0.104) |
| Aberrant motor behavior | 0.014 (-0.004,0.033) | 0.003 (-0.002,0.009) | -0.008 (-0.025,0.009) | 0.002 (-0.045,0.05) | 0.011 (-0.047,0.069) |
| Sleep/nightime behaviors | 0.005 (-0.015,0.025) | 0.002 (-0.004,0.008) | 0.001 (-0.017,0.019) | -0.018 (-0.069,0.035) | -0.010 (-0.074,0.053) |
| Appetite/eating disturbances | -0.014 (-0.035,0.006) | 0 (-0.006,0.006) | 0.007 (-0.01,0.024) | -0.025 (-0.075,0.027) | -0.031 (-0.093,0.030) |

* p<0.05
** p<0.01
*** p<0.001
+ p_FDR <0.05

**Discussion: Mediation Approach and Limitations**

We did not formally test the possibility that NPI items could influence cognition (i.e., reverse causality) because it was not within the pre-specified scope of our paper, though others have found such a potential directional association in a multinational study, as the presence of psychotic symptoms was associated with greater cognitive decline **^1^.** Nevertheless, we clearly demonstrated that cognition as a mediator could also causally influence a wide range of NPI items.

There are several limitations to our study. Because we examined postmortem data test results, NPI ratings were all measured relatively late in the disease process or ex vivo in the case of pathologies. However, we found that there was little difference in mean scores or last score when we examined this previously^2^**^3^.** Moreover, the types of physiologic changes that might be captured by direct e4 paths are often thought to be prominent late in the disease course and as such allowed us to test our hypothesis. Last, there is an ascertainment bias as the sample was collected in ADRCs whose charge was to study AD and a lack of ethnic diversity as the sample was largely White. The latter may reduce generalizability.

1.       Scarmeas, N.*, et al.* Delusions and hallucinations are associated with worse outcome in Alzheimer disease. *Arch Neurol* **62**, 1601-1608 (2005).

**Supplementary Figure 1. Legend. Flow Chart of the Derivation of the Sample. N=1199 for the Primary Analysis**

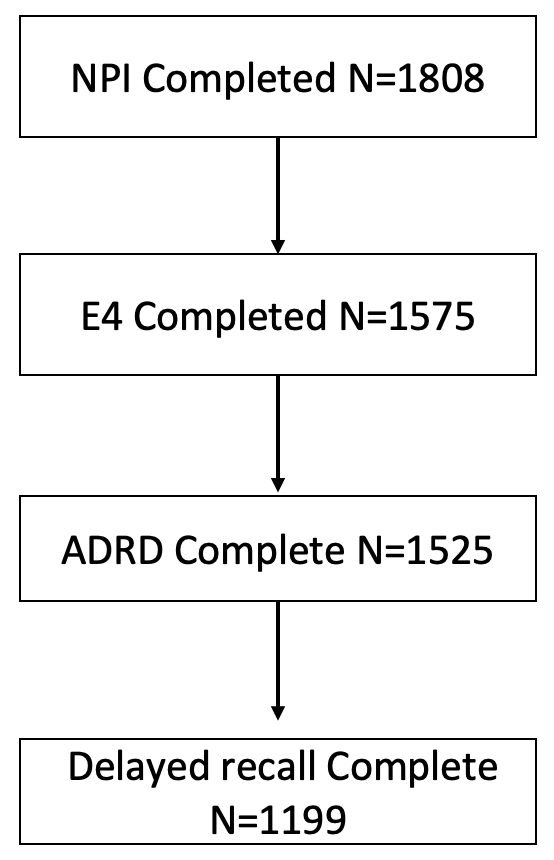
